# Polygenic risk score and statin relative risk reduction for primary prevention in a real-world population

**DOI:** 10.1101/2021.07.28.21254571

**Authors:** Akinyemi Oni-Orisan, Tanushree Haldar, Mari A. S. Cayabyab, Dilrini K. Ranatunga, Thomas J. Hoffmann, Carlos Iribarren, Ronald M. Krauss, Neil Risch

**Affiliations:** Department of Clinical Pharmacy, University of California San Francisco, San Francisco, California, USA; Institute for Human Genetics, University of California San Francisco, San Francisco, California, USA; Department of Bioengineering and Therapeutic Sciences, University of California San Francisco, San Francisco, California, USA; Kaiser Permanente Division of Research, Oakland, California, USA; Department of Epidemiology and Biostatistics, University of California San Francisco, San Francisco, California, USA; Department of Pediatrics, University of California San Francisco, Oakland, California, USA; Department of Medicine, University of California San Francisco, Oakland, Cali fornia, USA

**Keywords:** Coronary Heart Disease, Precision medicine, PRS

## Abstract

**Background:** Randomized-controlled trials demonstrate that high coronary heart disease (CHD) polygenic risk score modifies statin CHD relative risk reduction, but it is unknown if the association extends to statin users undergoing routine care.

**Objectives:** The primary objective was to determine how statin effectiveness is modified by CHD polygenic risk score in a real-world cohort of primary prevention participants.

**Methods:** We determined polygenic risk scores in participants of the Genetic Epidemiology Research on Adult Health and Aging (GERA) cohort. Cox regression models were used to compare the risk of the cardiovascular outcomes between statin users and matched nonusers.

**Results:** The hazard ratio (HR) for statin effectiveness on incident myocardial infarction was similar within 10-year atherosclerotic cardiovascular disease (ASCVD) risk score groups at 0.65 (95% confidence interval [CI] 0.39-1.08; *P*=0.10), 0.65 (95% CI 0.56-0.77; *P*=2.1E-7), and 0.67 (95% CI 0.57-0.80; *P*=4.3E-6) for borderline, intermediate, and high ASCVD groups, respectively. In contrast, statin effectiveness by polygenic risk was largest in the high polygenic risk score group (HR 0.62, 95% CI 0.50-0.77; *P*=1.4E-5), intermediate in the intermediate polygenic risk score group (HR 0.70, 95% CI 0.61-0.80; *P*=5.7E-7), and smallest in the low polygenic risk score group (HR 0.86, 95% CI 0.65-1.16; *P*=0.33). ASCVD risk and statin LDL-C lowering did not differ across polygenic risk score groups.

**Conclusions:** In primary prevention patients undergoing routine care, CHD polygenic risk modified statin relative risk reduction of incident myocardial infarction independent of statin LDL-C lowering. Our findings extend prior work by identifying a subset of patients with attenuated clinical benefit from statins.

## Introduction

Statin therapy has shown substantial benefit in preventing atherosclerotic cardiovascular disease (ASCVD) events including coronary heart disease (CHD) and other vascular events(1). The magnitude of this observed benefit was found to be directly proportional to the degree of statin-induced low-density lipoprotein cholesterol (LDL-C) lowering(1, 2). Although clinical and demographic factors (age, sex, diabetes, hypertension, etc.) were not found to modify this relationship(1, 2), the discovery of modifiers could improve current practice guidelines regarding eligibility for statin treatment.

Recent genetic substudies of statin randomized-controlled trials have demonstrated that factors other than the degree of LDL-C lowering can modify statin CHD relative risk reduction. In particular, high CHD polygenic risk (derived from variants previously found to be associated with CHD in genome-wide association studies(3–5)) was associated with a greater statin benefit compared to low and intermediate polygenic risk score groups (6, 7). In addition to statin-induced LDL-C lowering, this association was also found to be independent of other traditional ASCVD risk factors including strong family history.

Although these results suggest that polygenic risk scores may be able to better identify candidates predicted to receive substantial benefit from statin therapy beyond those who qualify for treatment based on current practice guidelines, results from randomized-controlled trials may not generalize to the broad population of statin users undergoing routine clinical care(8). Thus, it is unknown if the association between CHD polygenic risk and statin CHD relative risk reduction extends to statin users outside of a clinical trial setting. The primary objective of this study was to validate that this previously-established CHD polygenic risk score modifies statin CHD risk reduction in a cohort of real-world, primary prevention participants. Furthermore, results from the aforementioned studies (both the statin randomized-controlled trial substudies as well as the genome wide association studies that derived the polygenic risk scores) are based on participants almost entirely self-identified as White race or of European descent (depending on the study)(3–7). Therefore, we also explored how this polygenic risk score modifies statin effectiveness in participants with substantial proportions of sub-Saharan African, Native American, and East Asian ancestry.

## Methods

### Data Source

We used data from participants in Genetic Epidemiology Research on Adult Health and Aging (GERA), a resource of the Research Program on Genes, Environment and Health (RPGEH) within the Kaiser Permanente Northern California (KPNC) health system that links electronic health record, genome-wide variant, and demographic survey data (9, 10) as described in the Supplemental Methods.

Approval was obtained from Kaiser Permanente and University of California Institutional Review Boards. Participants gave written informed consent.

### Genotyping

To maximize coverage of genome-wide variants, study participants were previously genotyped on one of four Affymetrix Axiom arrays based on self-reported race/ethnicity(11). Imputation was performed to the 1000 Genomes Project (Phase I integrated release, March 2012, with August 2012 chromosome X update and singletons removed)(12). Individual genetic ancestry admixture proportions were generated using the same algorithm as previously reported(13), except using Admixture v1.3.0(14) (a faster implementation of the same algorithm); we refer readers to Banda et al. for further details on how race/ethnicity and genetic ancestry were characterized in participants(13).

### Phenotyping

A participant met the definition of a statin user if (1) they were considered to be adherent from the date of statin initiation to the date of event or censor, (2) they appeared to be a new initiator of statin therapy within the time frame of follow-up, (3) their last statin dispensing record window ended ≤30 days before the date of event or censor, and (4) their date of statin initiation occurred >30 days before the date of event or censor (time-to-event and censoring analyses described in more detail below). We used proportion of days covered to estimate adherence and set 80% as the adherence threshold(15). To reduce the likelihood of including participants who were not new initiators of statins, we defined a new initiator of statin therapy as a participant whose initial statin dispensing record was >6 months after entering KPNC membership(10).

A participant met the definition of a statin nonuser if they had no statin dispensing records before the date of event or censoring unless statin initiation was considered to be too recent to impact outcomes (i.e., ≤30 days before event or censoring).

To identify statin nonusers with similar ASCVD risk as statin users at index, we matched each user to two nonusers by age (within 3 years at a maximum), sex, cigarette smoking, diabetes, hypertension, and race/ethnicity as described in the Supplemental Methods.

To obtain a primary prevention population, we included participants with no evidence of myocardial infarction prior to index. This definition for primary prevention was based on a prior genetic substudy of a statin randomized-controlled trial that investigated the association between the same polygenic risk score and statin CHD benefit(6).

As the main complication of CHD, we prespecified first-time myocardial infarction (fatal + non-fatal) to be the primary outcome. A major adverse cardiovascular event (MACE) was defined to include a myocardial infarction (fatal + non-fatal), an ischemic stroke (fatal + non-fatal), or any other cardiovascular disease death and first-time MACE was prespecified as the secondary outcome, consistent with the known benefit of statin therapy on all-cause cardiovascular mortality(16, 17). We generated time-to-event (from index) data with right-censoring. Participants who did not experience a primary or secondary outcome were censored at the time of the last LDL-C measurement in their records or at age 90 (due to lack of detailed data in this subgroup as previously described(10)), whichever occurred first.

### CHD polygenic risk score

In order to validate previous results from substudies of randomized-controlled trials as the key objective of this study, we selected variants among a total of 67 that were independently associated with CHD (and corresponding odds ratios) in prior genome wide association studies from European descent participants, as described(6). See Supplemental Methods for more details.

### Data analysis

For multiethnic cohorts in which sample size between self-identified race/ethnicity groups are unbalanced (such as GERA), any result from analyses in the combined cohort will likely represent the largest sample size thereby masking findings of underrepresented groups. Furthermore, despite its limitations(18), self-identified race is considered a “master status variable”(19) capturing important environmental (e.g., structural racism) and non-environmental (e.g., self-identified Black individuals in the United States often have substantial genetic ancestry from West and Central Africa(20)) factors that could be lost if not accounted for. For these reasons, and in accord with the previously-mentioned objectives of the study, we categorized participants by self-identified race/ethnicity groups: participants were stratified into race/ethnicity groups before subsequent analyses. Mean genetic ancestry proportions were calculated and reported within each race/ethnicity group, similar to previous studies(13), to ensure that ancestry from underrepresented populations were appropriately included in the study. Race/ethnicity groups with <250 statin users were not included in the study population due to inadequate statistical power.

First, Cox proportional hazards regression models were generated for the matched statin nonusers alone to compare the primary and secondary outcomes across polygenic risk score groups. Model covariates included sex, age, hypertension, diabetes, and cigarette smoking status at the time of index date. The low polygenic risk score group served as the reference. Second, Cox proportional hazards regression was used for statin effectiveness to compare the risk of each prespecified cardiovascular outcome between statin users and matched nonusers. Statin effectiveness for each outcome was determined overall, within polygenic risk score groups, and within 10-year ASCVD risk groups (using the Pooled Cohort Equations).

We conducted a secondary analysis to again investigate the risk of incident myocardial infarction across polygenic risk score groups in statin nonusers alone; however, in this case, we accounted for sample size differences between self-identified White participants and each of the other race/ethnicity groups (the sample size of other race/ethnicity groups that met the criteria for study inclusion were significantly smaller than White participants) as described in the Supplemental Methods. The purpose of this analysis was to determine if sample size explains the differences in the association between polygenic risk and incident myocardial infraction across race/ethnicity groups.

All analyses were conducted using R software (R Foundation for Statistical Computing, version 3.5.1, https://www.R-project.org/).

## Results

### Clinical characteristics and statin effectiveness in self-identified White participants

There were 10,915 self-identified White statin users who met the criteria for study inclusion. Mean genetic ancestry proportions in White statin users were 96.5%, 0.6%, 0.7%, and 0.5% for European, sub-Saharan African, Native American, and East Asian ancestry, respectively. Clinical characteristics were similar between statin users and matched nonusers (Supplemental Table 2) at time of index. This includes factors that were not matched for including 10-year ASCVD risk (mean risk was 13.7% [median 12.0%] and 13.2% [median 10.5%] in statin users and nonusers, respectively) and body mass index (median body mass index was 27.6 and 26.6 kilograms per meter squared in statin users and nonusers, respectively). At a median follow-up of 7.0 years (8.7 and 6.1 years in statin users and nonusers, respectively), the primary outcome occurred 1,565 times (460 and 1,105 times in statin users and nonusers, respectively) and the secondary outcome occurred 2,871 times (770 and 2,101 times in statin users and nonusers, respectively). Statins were effective in lowering LDL-C from the time of index (44% lowering and 68 milligrams per deciliter absolute reduction) compared to statin nonusers (4% lowering and 5 milligrams per deciliter absolute reduction). Statins significantly reduced the primary outcome of myocardial infarction (hazard ratio 0.71, 95% confidence interval 0.64-0.79; *P*=1.3E-9; Supplemental Figure 1A) and secondary outcome of MACE (hazard ratio 0.67, 95% confidence interval 0.61-0.72; *P*=3.3E-21; Supplemental Figure 1B) compared to statin nonusers.

### Polygenic risk score and cardiovascular outcomes in self-identified White participants

Clinical characteristics at index were similar in statin nonusers across polygenic risk score groups (Table 1, Supplemental Table 3). Higher CHD polygenic risk was associated with higher risk of myocardial infarction (hazard ratio 1.96 per polygenic risk score standard deviation, 95% confidence interval 1.68-2.29; *P*=1.5E-17; Figure 1) and MACE (hazard ratio 1.53 per polygenic risk score standard deviation, 95% confidence interval 1.36-1.71; *P*=1.6E-12; Supplemental Figure 2) in a linear fashion.

**Table 1:**
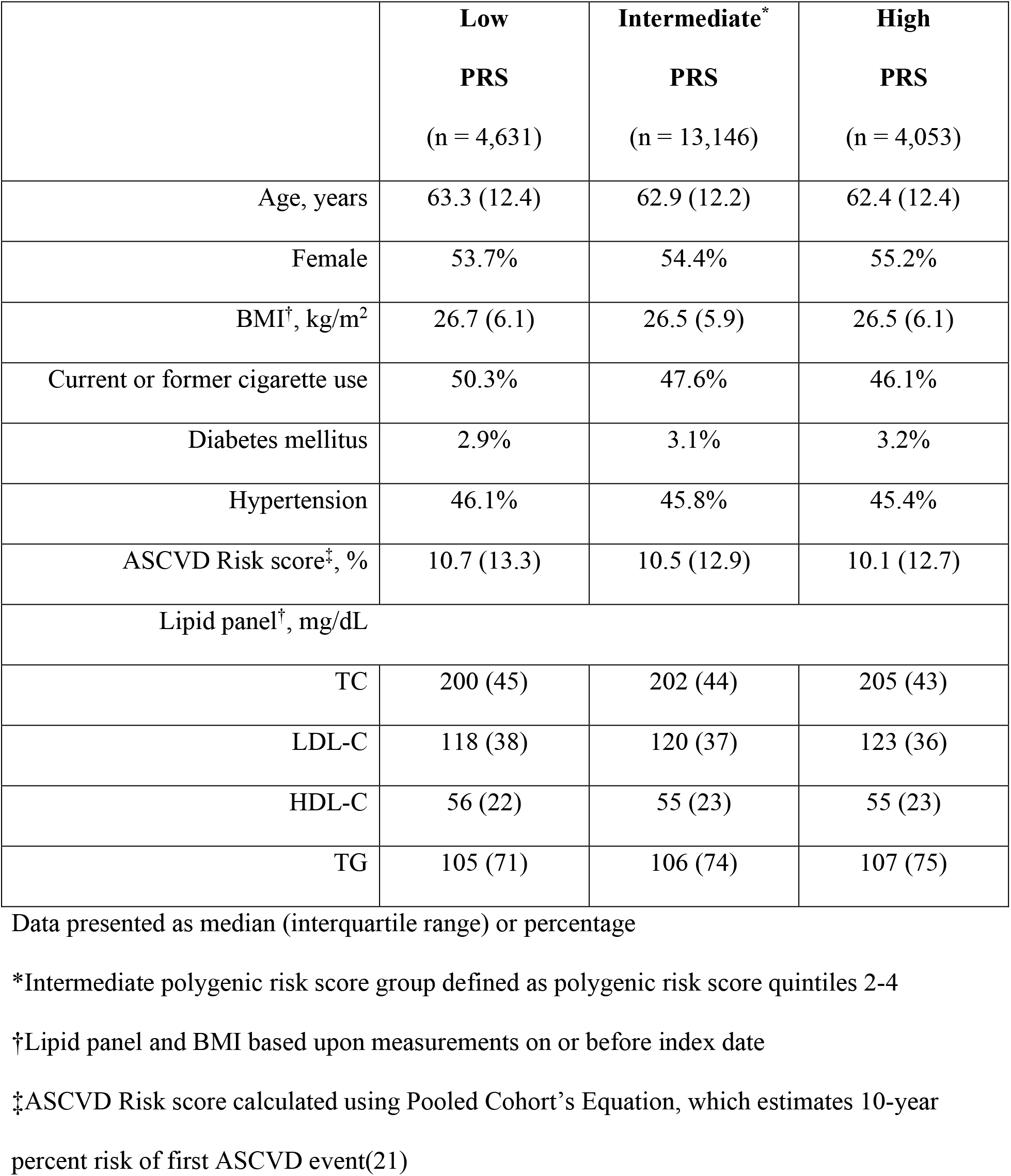
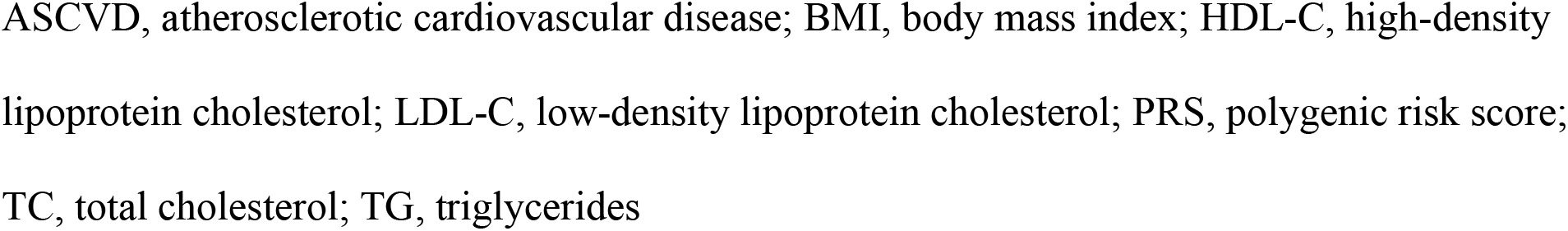
Clinical characteristics at index date across coronary heart disease polygenic risk score groups in self-identified White statin nonusers

|  | **Low**<br>**PRS**<br>**(n = 4,631)** | **Intermediate\***<br>**PRS**<br>**(n = 13,146)** | **High**<br>**PRS**<br>**(n = 4,053)** |
| --- | --- | --- | --- |
| Age, years | 63.3 (12.4) | 62.9 (12.2) | 62.4 (12.4) |
| Female | 53.7% | 54.4% | 55.2% |
| BMI<sup>†</sup>, kg/m<sup>2</sup> | 26.7 (6.1) | 26.5 (5.9) | 26.5 (6.1) |
| Current or former cigarette use | 50.3% | 47.6% | 46.1% |
| Diabetes mellitus | 2.9% | 3.1% | 3.2% |
| Hypertension | 46.1% | 45.8% | 45.4% |
| ASCVD Risk score<sup>‡</sup>, % | 10.7 (13.3) | 10.5 (12.9) | 10.1 (12.7) |
| Lipid panel<sup>†</sup>, mg/dL |  |  |  |
| TC | 200 (45) | 202 (44) | 205 (43) |
| LDL-C | 118 (38) | 120 (37) | 123 (36) |
| HDL-C | 56 (22) | 55 (23) | 55 (23) |
| TG | 105 (71) | 106 (74) | 107 (75) |

Data presented as median (interquartile range) or percentage
\*Intermediate polygenic risk score group defined as polygenic risk score quintiles 2-4
<sup>†</sup>Lipid panel and BMI based upon measurements on or before index date
<sup>‡</sup>ASCVD Risk score calculated using Pooled Cohort's Equation, which estimates 10-year percent risk of first ASCVD event(21)
ASCVD, atherosclerotic cardiovascular disease; BMI, body mass index; HDL-C, high-density lipoprotein cholesterol; LDL-C, low-density lipoprotein cholesterol; PRS, polygenic risk score; TC, total cholesterol; TG, triglycerides

**Figure 1.**
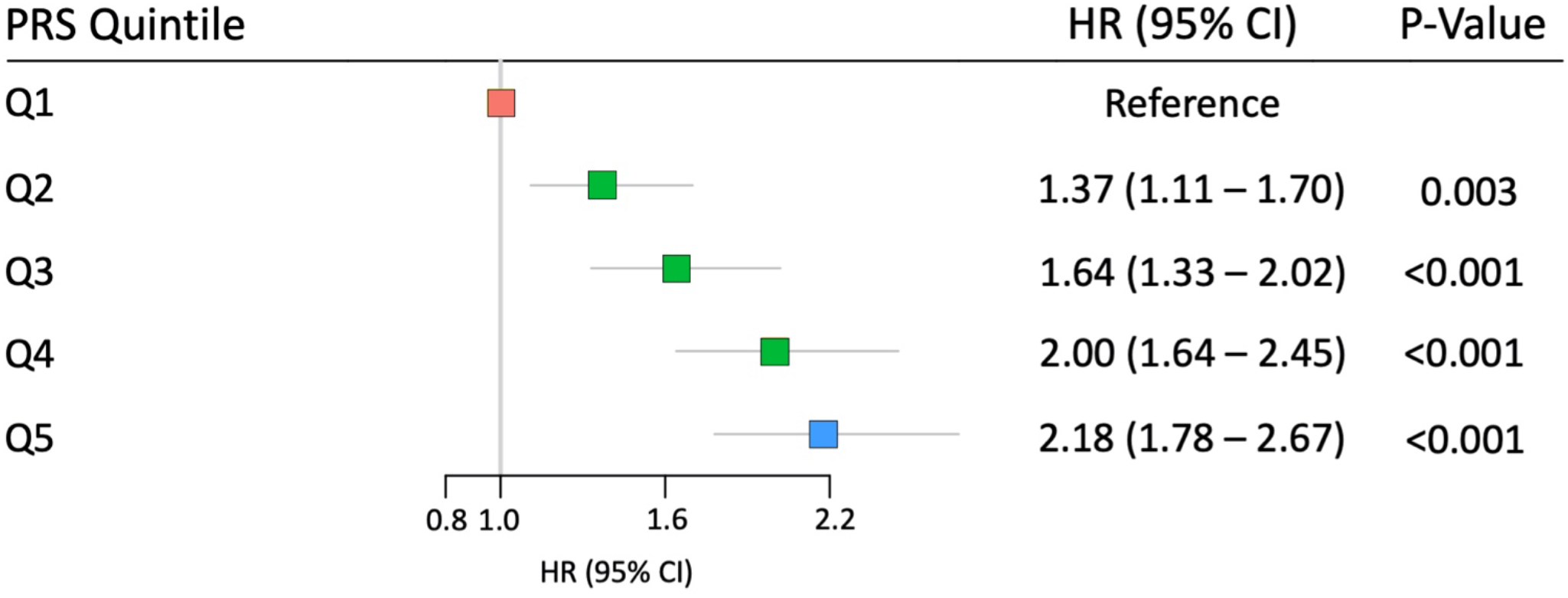
Risk of incident myocardial infarction across coronary heart disease polygenic risk score quintiles in self-identified White statin nonusers. Higher polygenic risk score was associated with an increasing risk gradient for incident myocardial infarction. CI, confidence interval; HR, hazard ratio; PRS, polygenic risk score

### Statin effectiveness by 10-year ASCVD risk score group in self-identified White participants

Within 10-year ASCVD risk score groups for which statin therapy may be considered based on national guidelines(21), the hazard ratio for statin effectiveness on incident myocardial infarction was 0.65 (95% confidence interval 0.39-1.08; *P*=0.10), 0.65 (95% confidence interval 0.56-0.77; *P*=2.1E-7), and 0.67 (95% confidence interval 0.57-0.80; *P*=4.3E-6) for borderline, intermediate, and high ASCVD risk, respectively (Figure 2A). For low ASCVD risk, which does not warrant statin therapy in primary prevention per national guidelines(21), the hazard ratio for statin effectiveness on incident myocardial infarction was 1.24 (95% confidence interval 0.79-1.92; *P*=0.35).

**Figure 2.**
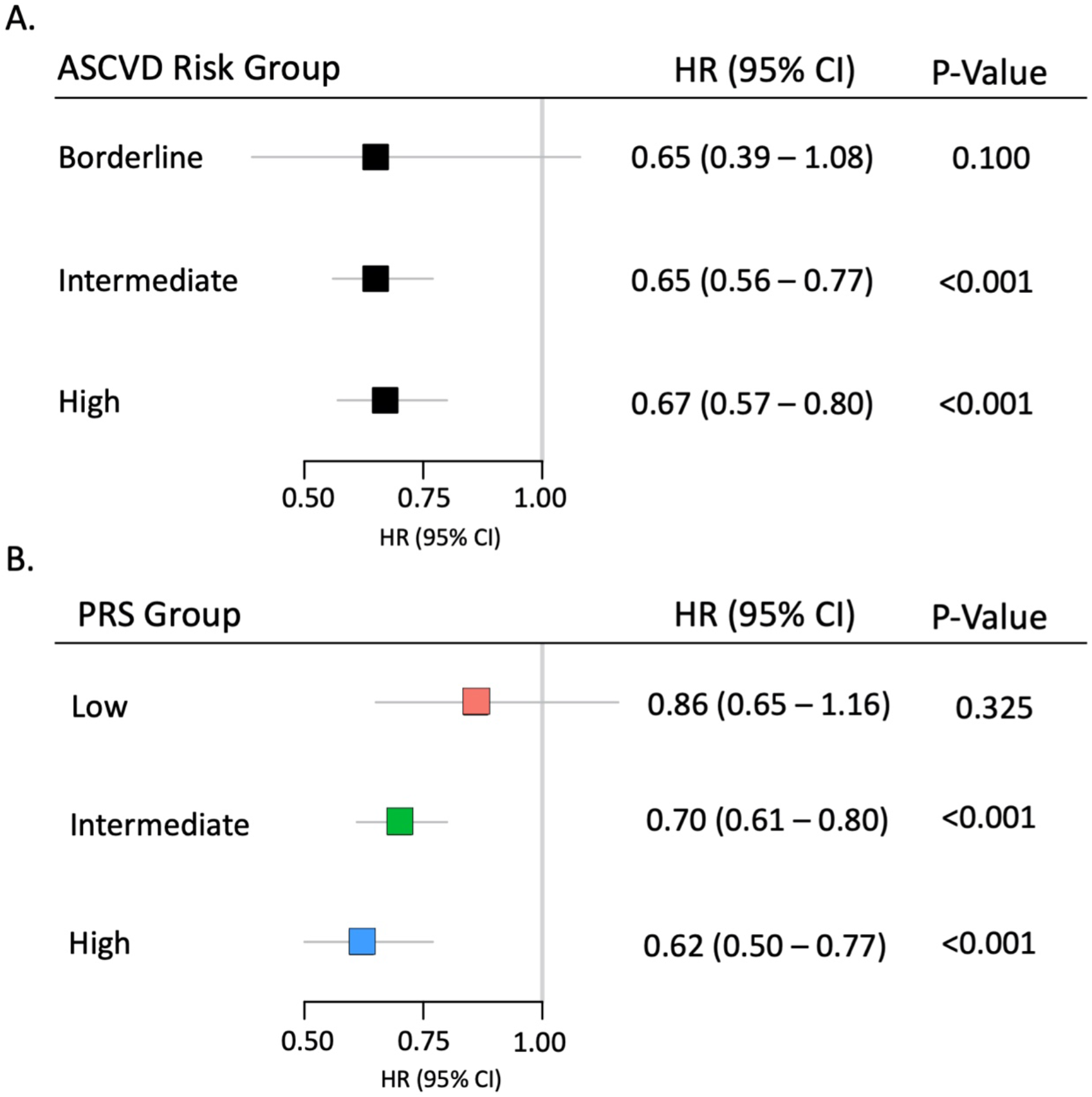
Statin effectiveness on incident myocardial infarction in self-identified White participants. Statin effectiveness did not vary across 10-year Pooled Cohort Equations ASCVD risk score groups in self-identified White participants (A). In contrast, statin effectiveness varied in these same participants across low (quintile 1), intermediate (quintiles 2-4), and high (quintile 5) polygenic risk score groups with smallest statin benefit in the low polygenic risk score group and largest benefit in the high polygenic risk score group (*P* for high versus low=0.04; B). ASCVD, atherosclerotic cardiovascular disease; CI, confidence interval; HR, hazard ratio; PRS, polygenic risk score

### Statin effectiveness by polygenic risk score group in self-identified White participants

On-treatment LDL-C and statin LDL-C lowering did not differ across polygenic risk score groups (Table 2; Supplemental Table 4). Statin effectiveness for the primary outcome was largest in the high polygenic risk score group (hazard ratio 0.62, 95% confidence interval 0.50-0.77; *P*=1.4E-5), middle in the intermediate polygenic risk score group (hazard ratio 0.70, 95% confidence interval 0.61-0.80; *P*=5.7E-7), and smallest in the low polygenic risk score group (hazard ratio 0.86, 95% confidence interval 0.65-1.16; *P*=0.33; *P* for high versus low=0.04; Central Illustration; Figure 2B; Table 2; Supplemental Table 4). Number-needed-to-treat corresponded to the pattern of statin relative risk reduction with values of 54, 79, and 217 for high, intermediate, and low polygenic risk score groups, respectively. We observed similar findings in statin effectiveness for MACE (Supplemental Figure 3; Table 2; Supplemental Table 4).

**Table 2:**
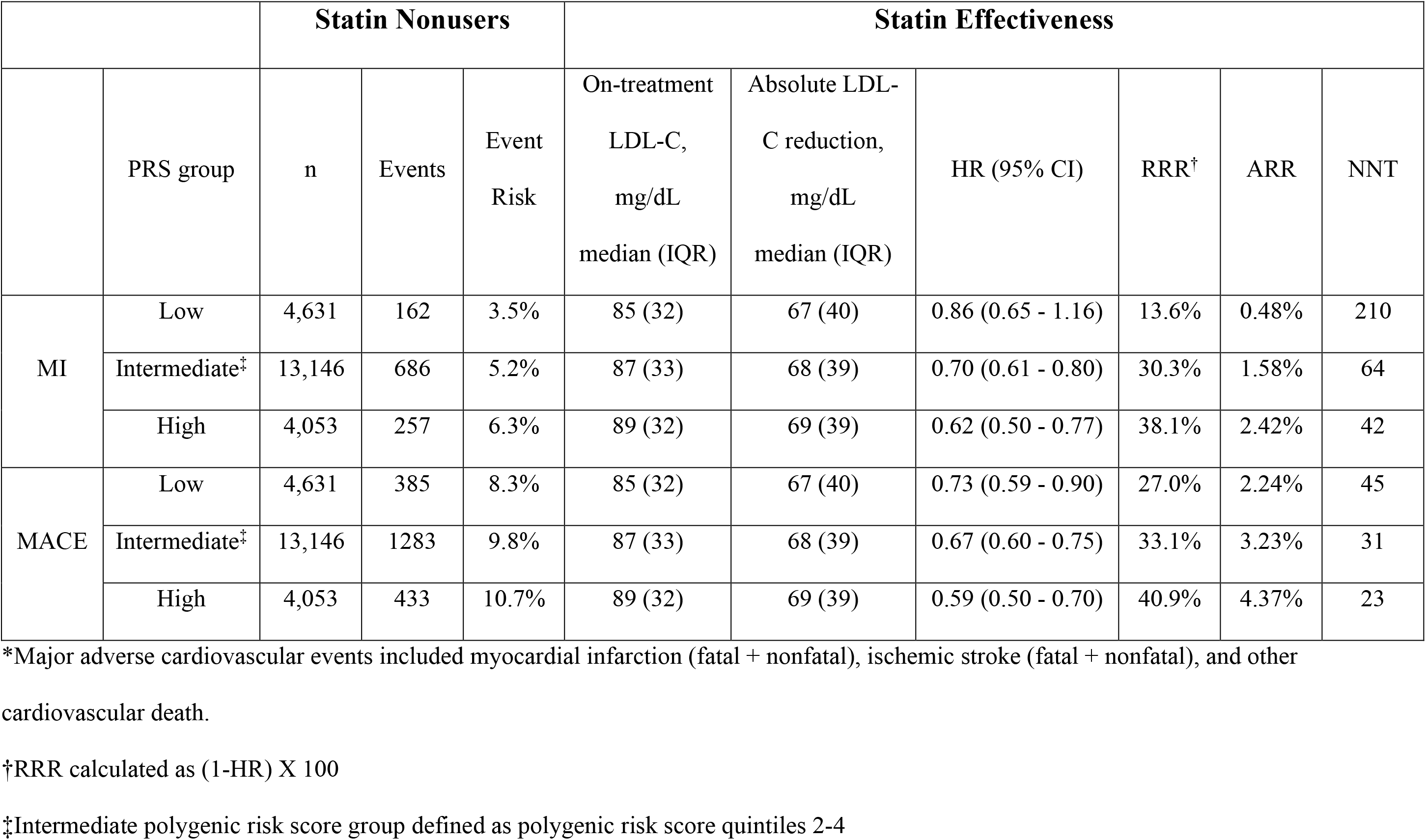
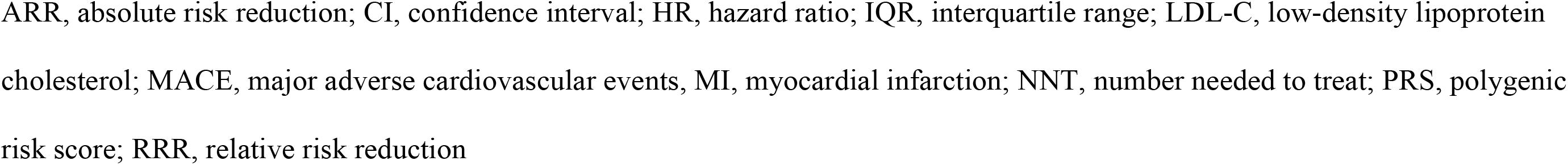
Statin effectiveness on incident myocardial infarction and major adverse cardiovascular events* across coronary heart disease polygenic risk score groups in self-identified White participants

|  |  | Statin Nonusers |  |  | Statin Effectiveness |  |  |  |  |  |
| --- | --- | --- | --- | --- | --- | --- | --- | --- | --- | --- |
|  | PRS group | n | Events | Event Risk | On-treatment LDL-C, mg/dL median (IQR) | Absolute LDL-C reduction, mg/dL median (IQR) | HR (95% CI) | RRR<sup>†</sup> | ARR | NNT |
| MI | Low | 4,631 | 162 | 3.5% | 85 (32) | 67 (40) | 0.86 (0.65 - 1.16) | 13.6% | 0.48% | 210 |
|  | Intermediate<sup>‡</sup> | 13,146 | 686 | 5.2% | 87 (33) | 68 (39) | 0.70 (0.61 - 0.80) | 30.3% | 1.58% | 64 |
|  | High | 4,053 | 257 | 6.3% | 89 (32) | 69 (39) | 0.62 (0.50 - 0.77) | 38.1% | 2.42% | 42 |
| MACE | Low | 4,631 | 385 | 8.3% | 85 (32) | 67 (40) | 0.73 (0.59 - 0.90) | 27.0% | 2.24% | 45 |
|  | Intermediate<sup>‡</sup> | 13,146 | 1283 | 9.8% | 87 (33) | 68 (39) | 0.67 (0.60 - 0.75) | 33.1% | 3.23% | 31 |
|  | High | 4,053 | 433 | 10.7% | 89 (32) | 69 (39) | 0.59 (0.50 - 0.70) | 40.9% | 4.37% | 23 |

\*Major adverse cardiovascular events included myocardial infarction (fatal + nonfatal), ischemic stroke (fatal + nonfatal), and other cardiovascular death.
<sup>†</sup>RRR calculated as (1-HR) X 100
<sup>‡</sup>Intermediate polygenic risk score group defined as polygenic risk score quintiles 2-4
ARR, absolute risk reduction; CI, confidence interval; HR, hazard ratio; IQR, interquartile range; LDL-C, low-density lipoprotein cholesterol; MACE, major adverse cardiovascular events, MI, myocardial infarction; NNT, number needed to treat; PRS, polygenic risk score; RRR, relative risk reduction

### Cardiovascular outcomes and statin effectiveness by polygenic risk score group in self-identified Black, Latinx, and East Asian participants

There were 300, 809, and 823 statin users meeting study inclusion criteria for self-identified Black, East Asian, and Latinx participants, respectively. There were not enough eligible statin users (<250) within each of the remaining race/ethnicity categories (Pacific Islander, South Asian, or Native American) for study inclusion. Mean genetic ancestry proportions were 73.5% sub-Saharan African, 1.4% East Asian, 0.2% Native American, and 22.7% European for Black; 0.2% sub-Saharan African, 93.9% East Asian, 0.2% Native American, and 4.1% European for East Asian; and 4.0% sub-Saharan African, 2.5% East Asian, 24.6% Native American, and 67.6% European for Latinx race/ethnicity groups. Clinical characteristics at the time of index and overall statin effectiveness results are presented in Supplemental Tables 5-7. Polygenic risk score distributions by race/ethnicity groups are shown in Supplemental Figure 4.

The relationship between polygenic risk score and the primary outcome of incident myocardial infarction in statin nonusers varied by race/ethnicity: the hazard ratio for this outcome was 1.47 (95% confidence interval 0.44-4.88; *P*=0.53), 2.35 (95% confidence interval 1.13-4.87; *P*=0.02), and 1.71 (95% confidence interval 0.94-3.13; *P*=0.08) per polygenic risk score standard deviation in Black, East Asian, and Latinx participants, respectively (Supplemental Figures 5-7).

When accounting for sample size differences, the beta (hazard ratio) characterizing the association between polygenic risk score and time-to-event for the primary outcome in statin nonusers did not deviate in Black, East Asian, or Latinx participants from the mean beta in White participants (Supplemental Figure 8). In contrast, the standard error characterizing the association between polygenic risk score and time-to-event for the primary outcome deviated furthest from the mean standard error of White statin nonusers in East Asian statin nonusers (4.27 standard deviations; *P*=4.0E-6). Deviation from the mean standard error of White statin nonusers was 2.14 (*P*=0.03) and 0.87 (*P*=0.18) standard deviations for the standard error characterizing the association between polygenic risk score and time-to-event in Black and Latinx statin nonusers, respectively (Supplemental Figure 9).

The relationship between polygenic risk score and statin effectiveness varied by race/ethnicity. In Black participants, the hazard ratio associated with statin use for the primary outcome was 0.65 (95% confidence interval 0.12-3.37; *P*=0.61), 0.70 (95% confidence interval 0.30-1.67; *P*=0.43), and 0.29 (95% confidence interval 0.03-2.52; *P*=0.26) in low, intermediate, and high polygenic risk score groups, respectively (*P* for high versus low=0.28; Supplemental Table 8). In East Asian participants, the hazard ratio associated with statin use was 1.72 (95% confidence interval 0.62-4.76; *P*=0.30), 0.71 (95% confidence interval 0.37-1.35; *P*=0.30), and 0.45 (95% confidence interval 0.16-1.25; *P*=0.13) in low, intermediate, and high polygenic risk score groups, respectively (*P* for high versus low=0.03; Supplemental Table 8). In Latinx participants, the hazard ratio associated with statin use was 0.49 (95% confidence interval 0.16-1.54; *P*=0.23), 0.76 (95% confidence interval 0.44-1.30; *P*=0.31), and 0.83 (95% confidence interval 0.34-2.01; *P*=0.68) in low, intermediate, and high polygenic risk score groups, respectively (*P* for high versus low=0.76; Supplemental Table 8).

## Discussion

Previous studies reported a strong association between CHD polygenic risk score and statin relative risk reduction of cardiovascular outcomes independent of statin-induced LDL-C lowering in primary prevention(6, 7). The polygenic risk score and its association with statin benefit was respectively generated from and studied in predominantly European descent participants(6, 7). The current study is the first to validates these results in a real-world cohort. Furthermore, this is the first study to demonstrate attenuated statin benefit in low polygenic risk score participants. Finally, this is the first to study this relationship in participants with substantial proportions of sub-Saharan African, Native American, and East Asian ancestry.

In our self-identified White participants, over a median follow-up of ∼7.0 years, we generated a statin response phenotype of 29% relative risk reduction for the primary outcome with a mean LDL-C lowering effect of 69 milligrams per deciliter in the overall population. Furthermore, we showed that this statin effect does not vary across the three guideline-based 10-year ASCVD risk score groups for which statin therapy should be at least considered for treatment (i.e., borderline, intermediate, and high ASCVD risk groups). These results are consistent with findings from meta-analyses of statin randomized-controlled trials, which reported a ∼23% relative risk reduction (in 5-year incidence of a major coronary event) for every 1 millimole per liter (39 milligrams per deciliter) of statin-induced LDL-C lowering and demonstrated that this statin risk reduction-cholesterol lowering relationship is not modified by traditional ASCVD risk factors. Our replication of these well-established statin effects illustrate the robustness of our phenotype as we moved forward in interrogating the impact of the CHD polygenic risk score.

We found that participants within the highest quintile of polygenic risk for CHD had >2-fold increased chance of incident myocardial infarction compared to the lowest quintile when not receiving statin therapy and an enhanced statin benefit (38% relative risk reduction for incident myocardial infarction) when receiving therapy. Genetic substudies of prior primary prevention randomized-controlled trials also demonstrate high baseline CHD risk and enhanced statin relative risk reduction in the highest polygenic risk score group(6, 7, 22). The WOSCOP substudy, for example, showed 1.66-fold enhanced risk of CHD in the placebo-treated participants and 44% statin relative risk reduction(6). Thus, our findings suggest that data from prior randomized-controlled trials can be extrapolated to patients undergoing routine care.

Participants in the lowest CHD polygenic risk score quintile received only a moderate statin relative risk reduction (14%), which was not found to be a statistically significant benefit compared to statin nonusers. This is a substantially weaker statin effect than for participants from the intermediate (30% risk reduction) and high polygenic risk score groups (38% risk reduction); these differences were observed despite an overall pretreatment risk profile (14% 10-year ASCVD risk) and statin LDL-C response (44% lowering from baseline) that did not differ by genetic background. In contrast, prior genetic randomized-controlled trial substudies in primary prevention populations did not find attenuated statin efficacy in the low polygenic risk score group compared to intermediate polygenic risk. For example, in a post-hoc analysis of the JUPITER [Justification for the Use of Statins in Prevention: An Intervention Trial Evaluating Rosuvastatin] trial, both low and intermediate polygenic risk participants experienced a statin relative risk reduction of 32% compared to placebo(7). Larger sample size and longer follow-up in the current study may have facilitated our elucidation of this association. Indeed, our sample size of 32,745 self-identified White, primary prevention participants is larger than the combined population of previous randomized-controlled trial genetic substudies investigating this topic. Additionally, the present real-world population may be more heterogenous (considering the strict inclusion/exclusion criteria of randomized-controlled trials), increasing the possibility of detecting this difference. For instance, elevated C-reactive protein and low LDL-C (<130 milligrams per deciliter) was required for enrollment in JUPITER(7), narrowing the generalizability of results from that trial. Overall, further studies are necessary to confirm this novel finding of the current study.

If replicated, the current results extend these prior data by demonstrating an additional potential clinical use of polygenic risk scores: to identify patients for whom statin therapy may not offer sufficient cardiovascular benefit (e.g., borderline ASCVD risk and low CHD polygenic risk score). The benefits of deprescribing (or never initiating) regardless of drug – in populations vulnerable to the impact of polypharmacy – continue to gain traction(23). Thus, there is a need for precision medicine tools that can optimize the risk and benefit of therapies. However, the current evidence must be interpreted with caution and followed up with future investigations before translation to the bedside.

We found variation in the relationship between polygenic risk score and both cardiovascular outcomes (statin nonusers alone) as well as statin risk relative reduction (statin users and nonusers) across populations. Unsurprisingly, the association between the CHD polygenic risk (derived from participants of European ancestry) and cardiovascular outcomes was strongest (i.e., smallest p-value) in our White participants. We conducted bootstrap analyses to determine the role of sample size in this observation: we measured the deviation among the other race/ethnicity groups from a distribution of parameters (betas and standard errors) characterizing the association between polygenic risk score and incident myocardial infarction in White participants (mean European ancestry of 96.5%). Our results show that even after accounting for sample size differences, variability (based on deviation from the distribution of standard errors) among other racial/ethnic populations was largest in those that had the least mean proportion of European ancestry (East Asian participants) and smallest in those with the most mean proportion of European ancestry (Latinx participants). This is congruent with prior data showing that polygenic risk scores may be most precise in the populations from which they were derived(24). Thus, it is of no surprise that the observed point estimates for statin relative risk reduction within each polygenic risk score group were also not consistent across populations.

### Study Limitations

These analyses in diverse populations are underpowered and exploratory in nature, but results may underscore the need for ancestry-specific CHD polygenic risk scores to advance statin precision medicine; efforts to generate CHD polygenic risk scores for minority populations are ongoing(25). The majority of human genomic research has been conducted in participants of European descent despite making up a relatively smaller proportion of the global population(24). Thus, although limited in sample numbers, our data in participants from underrepresented populations provide a much-needed foundation for future larger studies investigating this topic in statin users.

## Conclusion

In a large cohort of primary prevention patients undergoing routine care, CHD polygenic risk modified the effectiveness of statin therapy. Our findings (1) extend prior work by identifying a subset of patients less likely to receive clinical benefit from statins and (2) confirm results from genetic substudies of randomized controlled trials in high polygenic risk individuals, who receive enhanced statin benefit. More research is needed in larger, diverse cohorts to ensure that advances in polygenic risk scores will improve the health of all populations.

### Clinical perspectives

#### Competency in Systems-Based Practice

Incorporation of genome-wide data in the health care system to characterize a patient’s coronary heart disease polygenic risk score may facilitate risk/benefit analysis for statin therapy that may lead to improved clinical outcomes.

#### Translational Outlook #1

Future studies are needed that prospectively evaluate the clinical utility of polygenic risk score-guided statin therapy.

#### Competency in Medical Knowledge

Precision medicine tools developed from advances in genomics research may not benefit all patients if the study populations from which they are derived from are not fully inclusive.

#### Translational Outlook #2

Future studies in large, diverse populations are necessary to determine if genetic ancestry-specific polygenic risk scores improves the prediction of statin ASCVD benefit.

## Data Availability

https://www.ncbi.nlm.nih.gov/projects/gap/cgi-bin/study.cgi?study_id=phs000674.v2.p2

## Abbreviations

ASCVD: Atherosclerotic cardiovascular disease
CHD: coronary heart disease
LDL-C: low-density lipoprotein cholesterol
MACE: major adverse cardiovascular events

---

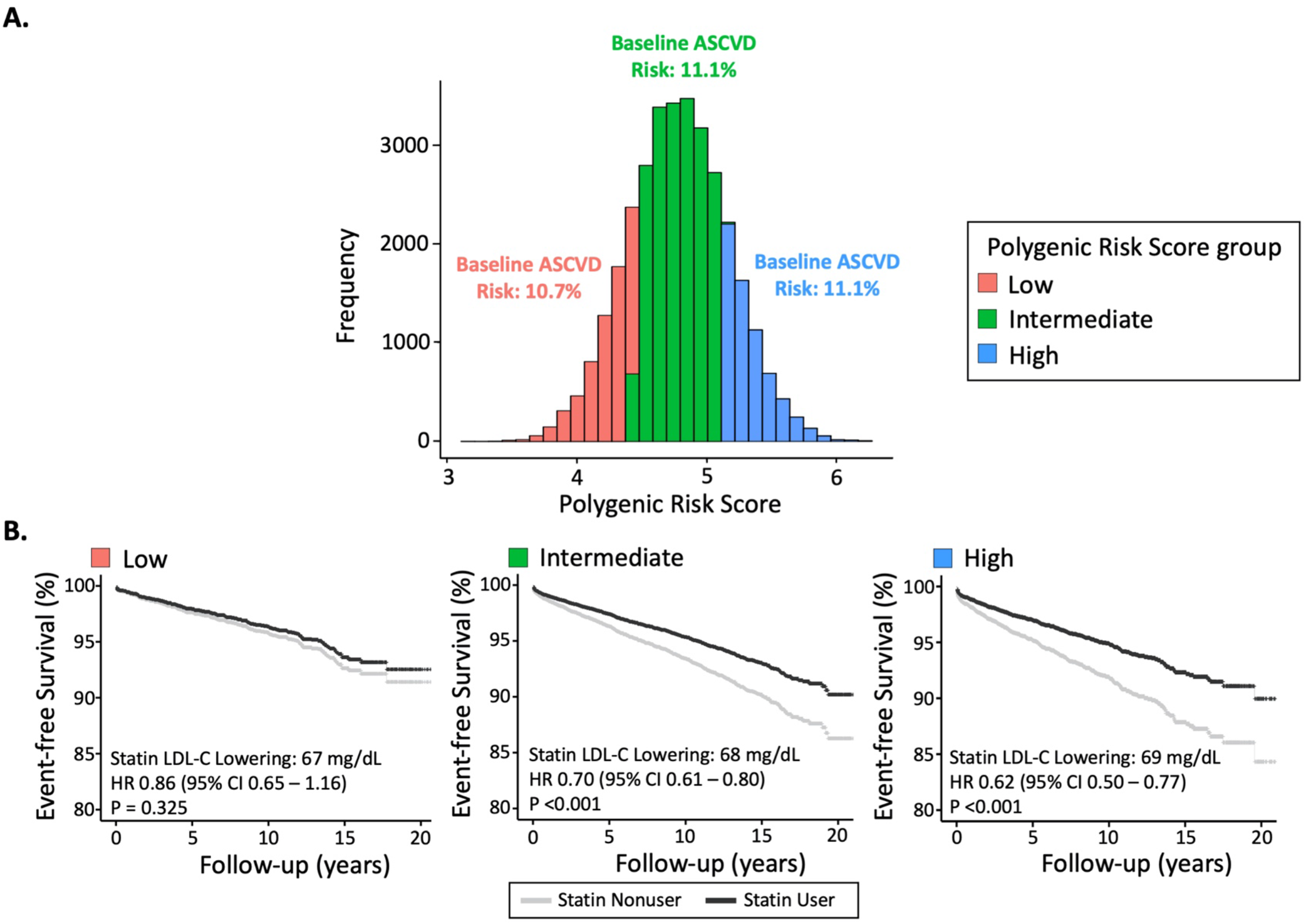
Central Illustration: Statin effectiveness on incident myocardial infarction by coronary heart disease polygenic risk score. Median 10-year ASCVD risk score for statin users and nonusers combined (using Pooled Cohort Equations) did not differ across low (quintile 1), intermediate (quintiles 2-4), and high (quintile 5) polygenic risk score groups at index date (A). Despite this similarity in baseline ASCVD risk by polygenic risk score group, statin effectiveness varied across groups with smallest benefit in the low polygenic risk score group and largest in the high polygenic risk score group (B). ASCVD, atherosclerotic cardiovascular disease; CI, confidence interval; HR, hazard ratio; LDL-C, low-density lipoprotein cholesterol

## Notes

### Competing Interest Statement

The authors have declared no competing interest.

### Funding Statement

This work was supported by grants RC2 AG036607, K01 HL143109, and P50 GM115318 from the National Institutes of Health (NIH). The development of the Research Program on Genes, Environment, and Health was supported by grants from the Robert Wood Johnson Foundation, the Wayne and Gladys Valley Foundation, the Ellison Medical Foundation, and Kaiser Permanente Community Benefit Programs.

