## Supplemental Methods, Tables, and Figures for "Polygenic risk score and statin relative risk reduction for primary prevention in a real-world population"

Data Source

Statin dispensing records, lipid panel measurements, International Classification of Diseases (ICD) codes, demographic information (i.e., sex and age), and total cardiovascular disease (e.g., myocardial infarction, stroke, heart failure) mortality data were extracted from electronic health records linked to GERA (between 1996 and early 2018). Participants were considered to have hypertension with >1 ICD9 code of 401.X on different dates. Diabetes status was defined based on the KPNC diabetes registry criteria using ICD codes, laboratory data, and dispensing data, as previously described(1). Myocardial infarction data were based on ICD9 codes of 410.X. Prior myocardial infarction data (to help differentiate primary prevention from secondary prevention participants) were based on ICD codes of 412.X. Acute ischemic stroke events were based on ICD9 codes of 433.X and 434.X. Race/ethnicity groups previously derived from 23 race/ethnicity/nationality categories (Black, East Asian, Latinx, White, Pacific Islander, South Asian, and Native American) as well as cigarette smoking status (current/former versus never) were extracted from self-report survey data(2, 3).

Matching

Since both statin users and nonusers were frequently referred for testing with a fasting lipid panel as part of routine KPNC screening efforts, we used the dates of lipid panel measurement as potential index dates for matching. For statin users, the last lipid panel date on or before statin initiation was set as the index timepoint for matching. For statin nonusers, the date of lipid panel measurement that minimized age difference between a user and matched nonusers was set as the index timepoint. To confirm that ASCVD risk was similar post matching, 10-year ASCVD risk at index age was determined in statin users and nonusers using the Pooled Cohort Equations (which uses age, sex, smoking, diabetes, race, systolic blood pressure, use of antihypertensive therapy, total cholesterol, and high density lipoprotein cholesterol to determine risk)(4). Individuals with 10-year ASCVD risk <5%, ≥5-<7.5%, ≥7.5%-<20, and ≥20% were considered to have low, borderline, intermediate, and high ASCVD risk, respectively(5).To simplify blood pressure and antihypertensive therapy effect for the purposes of 10-year ASCVD risk score calculation, systolic blood pressure was considered to be 125mmHg. This was similar to a prior study in the GERA cohort(6). To address uncertainties related to detecting hypertension treatment status in the electronic health records (e.g., nonadherence or partially adherent, resistant/refractory hypertension that remains uncontrolled, use of medications which alter blood pressure that are not indicated for hypertension, white coat hypertension, masked hypertension)(7), all positive hypertension diagnoses were considered to be treated. For these same reasons, hypertension diagnoses required two ICD9 codes on different days as stated above.

CHD polygenic risk score

Among these variants, 66 were directly genotyped or imputed in GERA participants and thus were carried forward to generate polygenic risk scores for this analysis (Supplemental Table 1). Within each race/ethnicity group, imputation quality for all variants was sufficient (minimum INFO score > 0.30) and commonly high (median INFO score > 0.99). The count of each variant allele (or allelic dosage for imputed variants) was weighted (log of the odds ratios) and summed to calculate a polygenic risk score for each participant. Participants were then ranked into quintiles based on score. Subsequently, participants were also classified into low (quintile 1), intermediate (quintiles 2-4), and high (quintile 5) polygenic risk score groups.

Secondary analysis accounting for sample size differences between race/ethnicity groups

For statin nonusers from each of the smaller sample size self-identified race/ethnicity groups, we selected a random subset of White statin nonusers so that the sample sizes between populations were identical(8). Cox proportional hazard models of time-to-event from index for incident myocardial infarction were then generated across polygenic risk score as a continuous variable within each of the new samples of White statin nonusers (corresponding to each of the race/ethnicity groups). Model covariates included sex, age, hypertension, diabetes, and cigarette smoking status at the time of index. This process was subsequently repeated 5,000 times (i.e., another random subset of White statin nonusers was selected among the full study population with sampling replacement and risk of the primary outcome was recalculated across polygenic risk score) for each of the subsets of White participants. Empirical distributions of two statistical parameters (beta and standard error, each log-transformed) describing the association between polygenic risk score and time-to-event across the iterations were calculated for each subset of White participants (each subset was based on a different sample size corresponding to the sample size of another race/ethnicity group) and the deviation from the mean distribution of each parameter for the corresponding single result from each other race/ethnicity group was determined (one-sided P-value). Deviation of observed standard error (smaller sample size race/ethnicity groups) from the empirical distribution of standard errors in White statin nonusers was used as a measure of goodness of fit for the polygenic risk score.

**Supplemental table 1: Variants^*^ associated with coronary heart disease and strength of association^†^ in genome-wide association studies**

| **Locus** | **Gene** | **SNP** | **Risk Allele** | **CHD OR**^†^ | **Frequency**^‡^ | **CH** | **Position** | **Allele Frequency: White**^§^  **n = 32,745** | **Allele Frequency: Black**^§^  **n = 900** | **Allele Frequency: East Asian**^§^  **n = 2,427** | **Allele Frequency:**  **Latinx**^§^  **n = 2,469** |
| --- | --- | --- | --- | --- | --- | --- | --- | --- | --- | --- | --- |
| 1p32.3 | *PCSK9* | rs11206510 | T | 1.06 | 0.88 | 1 | 55496039 | 0.82 | 0.84 | 0.95 | 0.85 |
| 1p32.2 | *PPAP2B* | rs17114036 | A | 1.11 | 0.83 | 1 | 56962821 | 0.91 | 0.86 | 0.95 | 0.92 |
| 1p13.3 | *SORT1* | rs602633 | C | 1.12 | 0.45 | 1 | 109821511 | 0.78 | 0.31 | 0.93 | 0.77 |
| 1q21.3 | *IL6R* | rs4845625 | T | 1.06 | 0.87 | 1 | 154422067 | 0.42 | 0.32 | 0.53 | 0.38 |
| 1q41 | *MIA3* | rs17464857 | T | 1.05 | 0.12 | 1 | 222762709 | 0.85 | 0.90 | 0.99 | 0.89 |
| 2p24.1 | *APOB* | rs515135 | G | 1.07 | NA | 2 | 21286057 | 0.81 | 0.55 | 0.90 | 0.79 |
| 2p21 | *ABCG5-ABCG8* | rs6544713 | T | 1.06 | 0.31 | 2 | 44073881 | 0.31 | 0.17 | 0.02 | 0.23 |
| 2p11.2 | *VAMP5-VAMP8-GGCX* | rs1561198 | A | 1.06 | 0.13 | 2 | 85809989 | 0.46 | 0.66 | 0.39 | 0.42 |
| 2q22.3 | *ZEB2-AC074093.1* | rs2252641 | G | 1.06 | 0.29 | 2 | 145801461 | 0.45 | 0.79 | 0.78 | 0.53 |
| 2q33.2 | *WDR12* | rs6725887 | C | 1.12 | 0.14 | 2 | 203745885 | 0.13 | 0.04 | 0.02 | 0.10 |
| 2q37.1 | *KCNJ13-GIGYF2* | rs1801251 | A | 1.05 | 0.19 | 2 | 233633460 | 0.35 | 0.26 | 0.36 | 0.29 |
| 3q22.3 | *MRAS* | rs9818870 | T | 1.07 | 0.15 | 3 | 138122122 | 0.16 | 0.08 | 0.04 | 0.10 |
| 4q12 | *REST-NOA1* | rs17087335 | T | 1.06 | 0.80 | 4 | 57838583 | 0.19 | 0.20 | 0.40 | 0.21 |
| 4q31.22 | *EDNRA* | rs1878406 | T | 1.1 | 0.74 | 4 | 148393664 | 0.14 | 0.18 | 0.23 | 0.15 |
| 4q32.1 | *GUCY1A3* | rs7692387 | G | 1.08 | 0.81 | 4 | 156635309 | 0.81 | 0.92 | 0.78 | 0.81 |
| 5q31.1 | *SLC22A4-SLC22A5* | rs273909 | C | 1.07 | 0.62 | 5 | 131667353 | 0.11 | 0.03 | 0.03 | 0.10 |
| 6p24.1 | *PHACTR1* | rs9369640 | A | 1.09 | 0.61 | 6 | 12901441 | 0.62 | 0.42 | 0.94 | 0.62 |
| 6p21.31 | *ANKS1A* | rs12205331 | C | 1.04 | 0.01 | 6 | 34898455 | 0.80 | 0.95 | 0.96 | 0.83 |
| 6p21.2 | *KCNK5* | rs10947789 | T | 1.07 | 0.70 | 6 | 39174922 | 0.76 | 0.92 | 0.80 | 0.74 |
| 6q23.2 | *TCF21* | rs12190287 | C | 1.07 | 0.19 | 6 | 134214525 | 0.63 | 0.88 | 0.56 | 0.58 |
| 6q25.3 | *LPA* | rs2048327 | G | 1.06 | 0.62 | 6 | 160863532 | 0.36 | 0.12 | 0.44 | 0.40 |
| 6q25.3 | *LPA* | rs3798220 | C | 1.28 | 0.01 | 6 | 160961137 | 0.02 | 0.01 | 0.09 | 0.14 |
| 6q26 | *PLG* | rs4252120 | T | 1.07 | 0.85 | 6 | 161143608 | 0.71 | 0.81 | 0.99 | 0.78 |
| 7p21.1 | *HDAC9* | rs2023938 | G | 1.08 | 0.55 | 7 | 19036775 | 0.10 | 0.17 | 0.01 | 0.08 |
| 7q22.3 | NA | rs12539895 | A | 1.08 | 0.47 | 7 | 107091849 | 0.21 | 0.06 | 0.13 | 0.18 |
| 7q32.2 | *ZC3HC1* | rs11556924 | C | 1.09 | 0.03 | 7 | 129663496 | 0.63 | 0.90 | 0.92 | 0.72 |
| 7q36.1 | *NOS3* | rs3918226 | T | 1.14 | 0.21 | 7 | 150690176 | 0.06 | 0.02 | 0.00 | 0.07 |
| 8p21.3 | *LPL* | rs264 | G | 1.11 | 0.43 | 8 | 19813180 | 0.85 | 0.86 | 0.79 | 0.84 |
| 8q24.13 | *TRIB1* | rs2954029 | A | 1.06 | 0.36 | 8 | 126490972 | 0.54 | 0.65 | 0.48 | 0.60 |
| 9p21.3 | *CDKN2A* | rs3217992 | A | 1.16 | 0.46 | 9 | 22003223 | 0.38 | 0.17 | 0.52 | 0.39 |
| 9p21.3 | *CDKN2A* | rs1333049 | C | 1.21 | 0.91 | 9 | 22125503 | 0.49 | 0.27 | 0.51 | 0.48 |
| 9q31.3 | *SVEP1* | rs111245230 | C | 1.14 | 0.52 | 9 | 113169775 | 0.04 | 0.01 | 0.00 | 0.03 |
| 9q34.2 | *ABO* | rs579459 | C | 1.07 | NA | 9 | 136154168 | 0.21 | 0.15 | 0.21 | 0.19 |
| 10p11.23 | *KIAA1462* | rs2505083 | C | 1.06 | 0.09 | 10 | 30335122 | 0.42 | 0.14 | 0.20 | 0.40 |
| 10q11.21 | *CXCL12* | rs2047009 | C | 1.05 | 0.39 | 10 | 44539913 | 0.51 | 0.19 | 0.50 | 0.38 |
| 10q11.21 | *CXCL12* | rs501120 | A | 1.07 | 0.17 | 10 | 44753867 | 0.86 | 0.62 | 0.70 | 0.78 |
| 10q23.31 | *LIPA* | rs11203042 | T | 1.04 | 0.46 | 10 | 90989109 | 0.44 | 0.38 | 0.53 | 0.47 |
| 10q23.31 | *LIPA* | rs2246833 | T | 1.06 | 0.31 | 10 | 91005854 | 0.34 | 0.40 | 0.33 | 0.41 |
| 10q24.32 | *CYP17A1* | rs12413409 | G | 1.1 | 0.77 | 10 | 104719096 | 0.91 | 0.95 | 0.76 | 0.85 |
| 11p15.4 | *SWAP70* | rs10840293 | A | 1.06 | 0.45 | 11 | 9751196 | 0.58 | 0.49 | 0.36 | 0.56 |
| 11p15.3 | NA | rs11042937 | T | 1.04 | 0.43 | 11 | 10745394 | 0.49 | 0.70 | 0.81 | 0.62 |
| 11q22.3 | *PDGF* | rs974819 | A | 1.07 | 0.79 | 11 | 103660567 | 0.30 | 0.49 | 0.60 | 0.27 |
| 11q23.3 | *APOA5-APOA1* | rs9326246 | C | 1.09 | 0.54 | 11 | 116611733 | 0.07 | 0.02 | 0.24 | 0.11 |
| 12p13.3 | *LRP1* | rs11172113 | C | 1.06 | NA | 12 | 57527283 | 0.40 | 0.42 | 0.22 | 0.44 |
| 12q24.12 | *SH2B3* | rs3184504 | T | 1.07 | 0.46 | 12 | 111884608 | 0.50 | 0.13 | 0.02 | 0.34 |
| 12p24.31 | *SCARB1* | rs11057830 | A | 1.08 | 0.71 | 12 | 125307053 | 0.15 | 0.18 | 0.09 | 0.13 |
| 13q12.3 | *FLT1* | rs9319428 | A | 1.06 | 0.55 | 13 | 28973621 | 0.30 | 0.31 | 0.47 | 0.23 |
| 13q34 | *COL4A1* | rs4773144 | G | 1.07 | 0.16 | 13 | 110960712 | 0.47 | 0.37 | 0.39 | 0.42 |
| 13q34 | *COL4A1* | rs9515203 | T | 1.08 | 0.24 | 13 | 111049623 | 0.74 | 0.75 | 0.89 | 0.73 |
| 14q32.2 | *HHIPL1* | rs2895811 | C | 1.06 | 0.97 | 14 | 100133942 | 0.41 | 0.28 | 0.27 | 0.34 |
| 15q22.33 | *SMAD3* | rs17293632 | C | 1.05 | 0.75 | 15 | 67442596 | 0.76 | 0.92 | 0.95 | 0.83 |
| 15q25.1 | *ADAMTS7* | rs7173743 | T | 1.07 | 0.13 | 15 | 79141784 | 0.54 | 0.53 | 0.48 | 0.54 |
| 15q26.1 | *MFGE8-ABHD2* | rs8042271 | G | 1.1 | 0.79 | 15 | 89574218 | 0.95 | 0.58 | 0.70 | 0.75 |
| 15q26.1 | *FURIN* | rs17514846 | A | 1.07 | 0.48 | 15 | 91416550 | 0.49 | 0.78 | 0.18 | 0.37 |
| 16q13 | *CETP* | rs247616 | C | 1.05 | 0.80 | 16 | 56989590 | 0.68 | 0.74 | 0.83 | 0.71 |
| 17p13.3 | *SMG6* | rs2281727 | C | 1.04 | 0.46 | 17 | 2117945 | 0.35 | 0.41 | 0.25 | 0.35 |
| 17p11.2 | *RASD1* | rs12936587 | G | 1.06 | 0.13 | 17 | 17543722 | 0.54 | 0.69 | 0.85 | 0.61 |
| 17q21.32 | *UBE2Z* | rs15563 | C | 1.04 | 0.35 | 17 | 47005193 | 0.53 | 0.18 | 0.68 | 0.46 |
| 17q23.2 | *BCAS3* | rs8080784 | C | 1.06 | 0.10 | 17 | 59017025 | 0.15 | 0.36 | 0.15 | 0.16 |
| 18q21.32 | *PMAIP1-MC4R* | rs663129 | A | 1.06 | 0.40 | 18 | 57838401 | 0.23 | 0.30 | 0.16 | 0.17 |
| 19p13.2 | *ANGPTL4* | rs116843064 | G | 1.16 | 0.85 | 19 | 8429323 | 0.98 | 0.99 | 1.00 | 0.98 |
| 19p13.2 | *LDLR* | rs1122608 | G | 1.1 | 0.49 | 19 | 11163601 | 0.76 | 0.92 | 0.89 | 0.82 |
| 19q13.32 | *APOE-APOC1* | rs2075650 | G | 1.11 | 0.28 | 19 | 45395619 | 0.14 | 0.12 | 0.11 | 0.11 |
| 19q13.32 | *APOE-APOC1* | rs445925 | C | 1.13 | 0.37 | 19 | 45415640 | 0.89 | 0.73 | 0.99 | 0.93 |
| 21q22.11 | *KCNE2* | rs9982601 | T | 1.13 | 0.56 | 21 | 35599128 | 0.13 | 0.24 | 0.01 | 0.10 |
| 22q11.23 | *POM121L9P-ADORA22B* | rs180803 | G | 1.02 | 0.91 | 22 | 24658858 | 0.99 | 0.91 | 0.92 | 0.99 |

*Previous genome wide association studies identified 67 SNP variants that were independently associated with CHD at the threshold for genome-wide significance(9). Participants were predominantly of European descent. The majority of these variants (66) were available in GERA-linked genetic data

(either directly or imputed) for the current study.

†The strength of association between each variant and coronary heart disease in previous genome wide association studies

‡Risk allele frequencies noted in 1000G phase 3 EUR samples.

§Allele frequencies categorized by self-identified race/ethnicity for all participants (statin users and nonusers) that met the criteria for inclusion in the current study.

CHD, coronary heart disease; CH, chromosome; GERA, Genetic Epidemiology Research on Adult Health and Aging; NA, not applicable; OR, odds ratio; SNP, single nucleotide polymorphism

**Supplemental Table 2: Clinical characteristics of self-identified White statin users and matched nonusers^*^ at index date**

|  | **Statin User**  (n = 10,915) | **Statin Nonuser**  (n = 21,830) |
| --- | --- | --- |
| Age, years | 62.6 (11.5) | 62.9 (12.3) |
| Female | 54.4% | 54.4% |
| BMI^†^, kg/m^2^ | 27.6 (6.4) | 26.6 (6.0) |
| Current of former cigarette use | 47.9% | 47.9% |
| Diabetes mellitus | 3.1% | 3.1% |
| Hypertension | 45.8% | 45.8% |
| ASCVD Risk score^‡^, % | 12.0 (12.1) | 10.5 (12.9) |
| Lipid panel^†^, mg/dL |  |  |
| TC | 244 (52) | 203 (44) |
| LDL-C | 158 (43) | 120 (37) |
| HDL-C | 52 (20) | 55 (23) |
| TG | 139 (97) | 106 (74) |

Data presented as median (interquartile range) or percentage.

*Statin users and nonusers were matched in a one user to two nonusers ratio by the following factors: age (within 3 years), sex, cigarette use, diabetes status, hypertension status

†Lipid panel and BMI based upon measurements on or before index date

‡ASCVD Risk score calculated using Pooled Cohort’s Equation, which estimates 10-year percent risk of first ASCVD event(4).

ASCVD, atherosclerotic cardiovascular disease; BMI, body mass index; HDL-C, high-density lipoprotein cholesterol; LDL-C, low-density lipoprotein cholesterol; TC, total cholesterol; TG, triglycerides.

**Supplemental Table 3: Clinical characteristics at index date across coronary heart disease polygenic risk score quintiles in self-identified White statin nonusers**

|  | **Quintile 1**  (n = 4,631) | **Quintile 2**  (n = 4,486) | **Quintile 3**  (n = 4,375) | **Quintile 4**  (n = 4,285) | **Quintile 5**  (n = 4,053) |
| --- | --- | --- | --- | --- | --- |
| Age, years | 63.3 (12.4) | 63.1 (12.4) | 62.7 (12.1) | 62.9 (12.1) | 62.4 (12.4) |
| Sex (female) | 53.7% | 54.8% | 54.2% | 54.1% | 55.2% |
| BMI^*^, kg/m^2^ | 26.7 (6.1) | 26.6 (5.7) | 26.5 (5.9) | 26.5 (6.1) | 26.5 (6.1) |
| Cigarette use (current or former) | 50.3% | 48.8% | 46.7% | 47.1% | 46.1% |
| Diabetes mellitus | 2.9% | 3.4% | 2.8% | 3.2% | 3.2% |
| Hypertension | 46.1% | 46.3% | 45.6% | 45.5% | 45.4% |
| ASCVD Risk score^†^, % | 10.7 (13.3) | 10.6 (13.0) | 10.2 (12.4) | 10.5 (13.0) | 10.1 (12.7) |
| Lipid panel**^*^**, mg/dL |  |  |  |  |  |
| TC | 200 (45) | 201 (44) | 203 (44) | 203 (44) | 205 (43) |
| LDL-C | 118 (38) | 119 (37) | 121 (37) | 121 (37) | 123 (36) |
| HDL-C | 56 (22) | 55 (23) | 55 (22) | 55 (23) | 55 (23) |
| TG | 105 (71) | 106 (74) | 106 (75) | 106 (75) | 107 (75) |

Data presented as median (interquartile range) or percentage.

*Lipid panel and BMI based upon measurements on or before index date

†ASCVD Risk score calculated using Pooled Cohort’s Equation, which estimates 10-year percent risk of first ASCVD event(4).

ASCVD, atherosclerotic cardiovascular disease; BMI, body mass index; HDL-C, high-density lipoprotein cholesterol; LDL-C, low-density lipoprotein cholesterol; TC, total cholesterol; TG, triglycerides.

**Supplemental Table 4: Statin effectiveness on incident myocardial infarction and major adverse cardiovascular events* across coronary heart disease polygenic risk score groups in self-identified White participants**

|  | | **Statin Nonusers** | | | **Statin Effectiveness** | | | | | |
| --- | --- | --- | --- | --- | --- | --- | --- | --- | --- | --- |
|  | PRS Quintile | n | Events | Event Risk | On-treatment LDL-C, mg/dL  median (IQR) | Absolute LDL-C reduction, mg/dL median (IQR) | HR (95% CI) | RRR* | ARR | NNT |
| MI | Q1 | 4,631 | 162 | 3.5% | 85 (32) | 67 (40) | 0.86 (0.65 - 1.16) | 13.6% | 0.48% | 210 |
|  | Q2 | 4,486 | 203 | 4.5% | 88 (32) | 66 (39) | 0.65 (0.49 - 0.86) | 35.0% | 1.59% | 64 |
|  | Q3 | 4,375 | 220 | 5.0% | 87 (33) | 67 (38) | 0.71 (0.55 - 0.91) | 29.1% | 1.46% | 69 |
|  | Q4 | 4,285 | 263 | 6.1% | 87 (33) | 69 (40) | 0.70 (0.56 - 0.87) | 30.0% | 1.84% | 55 |
|  | Q5 | 4,053 | 257 | 6.3% | 89 (32) | 69 (39) | 0.62 (0.50 - 0.77) | 38.1% | 2.42% | 42 |
| MACE | Q1 | 4,631 | 385 | 8.3% | 85 (32) | 67 (40) | 0.73 (0.59 - 0.90) | 27.0% | 2.24% | 45 |
|  | Q2 | 4,486 | 417 | 9.3% | 88 (32) | 66 (39) | 0.62 (0.50 - 0.76) | 38.2% | 3.55% | 29 |
|  | Q3 | 4,375 | 415 | 9.5% | 87 (33) | 67 (38) | 0.72 (0.60 - 0.87) | 27.6% | 2.61% | 39 |
|  | Q4 | 4,285 | 451 | 10.5% | 87 (33) | 69 (40) | 0.65 (0.54 - 0.78) | 35.1% | 3.69% | 28 |
|  | Q5 | 4,053 | 433 | 10.7% | 89 (32) | 69 (39) | 0.59 (0.50 - 0.70) | 40.9% | 4.37% | 23 |

*Major adverse cardiovascular events include myocardial infarction (fatal + nonfatal), ischemic stroke (fatal + nonfatal), and other cardiovascular death.

†RRR calculated as (1-HR) X 100

ARR, absolute risk reduction; CI, confidence interval; HR, hazard ratio; IQR, interquartile range; LDL-C, low-density lipoprotein cholesterol; MACE, major adverse cardiovascular events; MI, myocardial infarction; NNT, number needed to treat; PRS, polygenic risk score; RRR, relative risk reduction

**Supplemental Table 5. Clinical characteristics of statin users and matched nonusers* by self-identified race/ethnicity**

|  | **Statin User** | **Statin Nonusers** |
| --- | --- | --- |
| Black | | |
| n | 300 | 600 |
| Age, years | 61.5 (10.4) | 61.7 (10.7) |
| Female | 57.3% | 57.3% |
| BMI^†^, kg/m^2^ | 30.2 (8.0) | 29.2 (6.7) |
| Current or former cigarette use | 48.7% | 48.7% |
| Diabetes mellitus | 7.3% | 7.3% |
| Hypertension | 73.3% | 73.3% |
| ASCVD Risk score^‡^, % | 13.9 (12.5) | 12.5 (13.4) |
| Lipid panel^†^, mg/dL |  |  |
| TC | 243 (55) | 196 (46) |
| LDL-C | 162 (49) | 118 (40) |
| HDL-C | 53 (18) | 56 (22) |
| TG | 120 (77) | 90 (55) |
| East Asian | | |
| n | 809 | 1,618 |
| Age, years | 60.0 (12.3) | 60.2 (12.6) |
| Female | 50.7% | 50.7% |
| BMI^†^, kg/m^2^ | 25.9 (4.5) | 24.5 (5.2) |
| Current or former cigarette use | 32.4% | 32.4% |
| Diabetes mellitus | 2.8% | 2.8% |
| Hypertension | 52.4% | 52.4% |
| ASCVD Risk score^‡^, % | 8.7 (10.8) | 7.7 (11.1) |
| Lipid panel^†^, mg/dL |  |  |
| TC | 246 (48) | 205 (38) |
| LDL-C | 158 (42) | 119 (36) |
| HDL-C | 53 (17) | 55 (23) |
| TG | 142 (101) | 115 (88) |
| Latinx | | |
| n | 823 | 1,646 |
| Age, years | 61.1 (11.8) | 61.1 (11.9) |
| Female | 54.2% | 54.2%% |
| BMI^†^, kg/m^2^ | 28.7 (6.2) | 28.1 (6.5) |
| Current or former cigarette use | 44.3% | 44.3% |
| Diabetes mellitus | 7.2% | 7.2% |
| Hypertension | 47.4% | 47.4% |
| ASCVD Risk score^‡^, % | 10.5 (12.5) | 9.2 (12.1) |
| Lipid panel^†^, mg/dL |  |  |
| TC | 243 (54) | 200 (43) |
| LDL-C | 156 (45) | 120 (36) |
| HDL-C | 49 (17) | 52 (20) |
| TG | 155 (92) | 121 (80) |

Data presented as median (interquartile range) or percentage

*Statin users and nonusers were matched in a one user to two nonusers ratio by the following factors: age (within 3 years at a maximum), sex, cigarette use, diabetes status, hypertension status

†Lipid panel and BMI based upon measurements on or before index date

‡ASCVD Risk score calculated using Pooled Cohort’s Equation, which estimates 10-year percent risk of first ASCVD event(4)

ASCVD, atherosclerotic cardiovascular disease; BMI, body mass index; HDL-C, high-density lipoprotein cholesterol; LDL-C, low-density lipoprotein cholesterol; TC, total cholesterol; TG, triglycerides

**Supplemental Table 6: Clinical characteristics at index date in statin nonusers across coronary heart disease polygenic risk score groups by self-identified race/ethnicity**

|  | **Low**  **PRS** | **Intermediate^*^ PRS** | **High**  **PRS** |
| --- | --- | --- | --- |
| Black | | | |
| n | 129 | 354 | 117 |
| Age, years | 61.3 (9.3) | 62.0 (11.3) | 61.6 (9.8) |
| Female | 58.1% | 55.9% | 60.7% |
| BMI^‡^, kg/m^2^ | 29.2 (7.3) | 29.5 (6.7) | 28.7 (5.2) |
| Current or former cigarette use | 55.0% | 46.6% | 47.9% |
| Diabetes mellitus | 6.2% | 7.3% | 8.5% |
| Hypertension | 72.9% | 74.3% | 70.9% |
| ASCVD Risk score^‡^, % | 13.8 (16.2) | 12.8 (11.9) | 11.0 (12.0) |
| Lipid panel^†^, mg/dL |  |  |  |
| TC | 187 (46) | 198 (43) | 199 (54) |
| LDL-C | 113 (38) | 119 (38) | 118 (43) |
| HDL-C | 55 (22) | 55 (21) | 58 (21) |
| TG | 84 (53) | 90 (54) | 99 (57) |
| East Asian | | | |
| n | 345 | 963 | 310 |
| Age, years | 60.0 (12.6) | 60.3 (12.3) | 60.2 (12.7) |
| Female | 51.0% | 49.8% | 52.9% |
| BMI^‡^, kg/m^2^ | 24.5 (4.8) | 24.7 (5.2) | 24.3 (5.4) |
| Current or former cigarette use | 27.8% | 34.1% | 32.3% |
| Diabetes mellitus | 3.8% | 2.4% | 3.2% |
| Hypertension | 51.9% | 53.3% | 50.3% |
| ASCVD Risk score^‡^, % | 7.2 (10.2) | 8.0 (11.5) | 7.8 (10.8) |
| Lipid panel^†^, mg/dL |  |  |  |
| TC | 205 (42) | 206 (37) | 203 (36) |
| LDL-C | 117 (38) | 119 (37) | 120 (30) |
| HDL-C | 57 (24) | 55 (21) | 55 (23) |
| TG | 113 (92) | 114 (87) | 116 (87) |
| Latinx | | | |
| n | 332 | 991 | 323 |
| Age, years | 61.6 (11.9) | 61.2 (11.5) | 60.9 (12.5) |
| Female | 53.0% | 54.8% | 53.6% |
| BMI^‡^, kg/m^2^ | 28.1 (6.4) | 27.9 (6.2) | 28.9 (7.5) |
| Current or former cigarette use | 43.1% | 44.5% | 45.2% |
| Diabetes mellitus | 6.0% | 7.2% | 8.4% |
| Hypertension | 47.6% | 45.0% | 54.5% |
| ASCVD Risk score^‡^, % | 9.0 (12.9) | 9.2 (12.1) | 9.3 (11.7) |
| Lipid panel^†^, mg/dL |  |  |  |
| TC | 197 (39) | 200 (44) | 206 (41) |
| LDL-C | 118 (35) | 120 (37) | 124 (40) |
| HDL-C | 52 (21) | 52 (20) | 51 (18) |
| TG | 111 (71) | 121 (83) | 132 (80) |

Data presented as median (interquartile range) or percentage

* Intermediate polygenic risk score group defined as polygenic risk quintiles 2-4

†Lipid panel and BMI based upon measurements on or before index date

‡ ASCVD Risk score calculated using Pooled Cohort’s Equation, which estimates 10-year percent risk of first ASCVD event(4)

ASCVD, atherosclerotic cardiovascular disease; BMI, body mass index; HDL-C, high-density lipoprotein cholesterol; LDL-C, low-density lipoprotein cholesterol; PRS, polygenic risk score; TC, total cholesterol; and TG, triglycerides

**Supplemental Table 7: Statin effectiveness on incident myocardial infarction and major adverse cardiovascular events^*^ by self-identified race/ethnicity group**

|  | **Statin Nonusers** | | | **Statin Effectiveness** | | | | | |
| --- | --- | --- | --- | --- | --- | --- | --- | --- | --- |
|  | n | Events | Event risk | On-treatment LDL-C, mg/dL  median (IQR) | Absolute LDL-C reduction, mg/dL median (IQR) | HR (95% CI) | RRR^†^ | ARR | NNT |
| Black | | | | | | | | | |
| MI | 600 | 26 | 4.3% | 85 (36) | 70 (45) | 0.61 (0.30 - 1.24) | 39.0% | 1.69% | 60 |
| MACE | 600 | 52 | 8.7% | 85 (36) | 70 (45) | 0.47 (0.27 - 0.81) | 53.0% | 4.59% | 22 |
| East Asian | | | | | | | | | |
| MI | 1,618 | 55 | 3.4% | 86 (32) | 70 (40) | 0.76 (0.47 - 1.23) | 24.0% | 0.82% | 123 |
| MACE | 1,618 | 91 | 5.6% | 86 (32) | 70 (40) | 0.67 (0.45 - 0.99) | 33.0% | 1.86% | 54 |
| Latinx | | | | | | | | | |
| MI | 1,646 | 65 | 4.0% | 81 (35) | 70 (42) | 0.73 (0.48 - 1.12) | 27.0% | 1.07% | 94 |
| MACE | 1,646 | 110 | 6.7% | 81 (35) | 70 (42) | 0.71 (0.50 - 0.99) | 29.0% | 1.94% | 52 |

*Major adverse cardiovascular events include myocardial infarction (fatal + nonfatal), ischemic stroke (fatal + nonfatal), and other cardiovascular death

†RRR calculated as (1-HR) X 100

ARR, absolute risk reduction; CI, confidence interval; HR, hazard ratio; IQR, interquartile range; LDL-C, low-density lipoprotein cholesterol; MACE, major adverse cardiovascular events; MI, myocardial infarction; NNT, number needed to treat; PRS, polygenic risk score; RRR, relative risk reduction

**Supplemental Table 8: Statin effectiveness on incident myocardial infarction and major adverse cardiovascular events^*^ across coronary heart disease polygenic risk score groups by self-identified race/ethnicity**

|  | | **Statin Nonusers** | | | **Statin Effectiveness** | | | | | |
| --- | --- | --- | --- | --- | --- | --- | --- | --- | --- | --- |
|  | PRS group | n | # Events | Event risk | On-treatment LDL-C, mg/dL  median (IQR) | Absolute LDL-C reduction, mg/dL median (IQR) | HR (95% CI) | RRR^†^ | ARR | NNT |
| Black | | | | | | | | | | |
| MI | Low | 129 | 5 | 3.9% | 90 (47) | 75 (58) | 0.65 (0.12 - 3.37) | 35.3% | 1.37% | 74 |
|  | Intermediate^‡^ | 354 | 16 | 4.5% | 85 (34) | 68 (38) | 0.70 (0.30 - 1.67) | 29.5% | 1.33% | 75 |
|  | High | 117 | 5 | 4.3% | 82 (33) | 74 (54) | 0.29 (0.03 - 2.52) | 70.7% | 3.02% | 34 |
| MACE | Low | 129 | 11 | 8.5% | 90 (47) | 75 (58) | 0.58 (0.18 - 1.83) | 42.2% | 3.59% | 27 |
|  | Intermediate^‡^ | 354 | 32 | 9.0% | 85 (34) | 68 (38) | 0.51 (0.26 - 1.01) | 48.8% | 4.41% | 23 |
|  | High | 117 | 9 | 7.7% | 82 (33) | 74 (54) | 0.27 (0.06 - 1.25) | 73.4% | 5.65% | 18 |
| East Asian | | | | | | | | | | |
| MI | Low | 345 | 8 | 2.3% | 88 (40) | 71 (43) | 1.72 (0.62 - 4.76) | NA | NA | NA |
|  | Intermediate^‡^ | 963 | 31 | 3.2% | 85 (31) | 71 (39) | 0.71 (0.37 - 1.35) | 29.4% | 0.95% | 106 |
|  | High | 310 | 16 | 5.2% | 88 (29) | 65 (41) | 0.45 (0.16 - 1.25) | 54.8% | 2.83% | 36 |
| MACE | Low | 345 | 20 | 5.8% | 88 (40) | 71 (43) | 0.85 (0.37 - 1.96) | 15.1% | 0.87% | 115 |
|  | Intermediate^‡^ | 963 | 50 | 5.2% | 85 (31) | 71 (39) | 0.78 (0.47 - 1.29) | 22.0% | 1.14% | 88 |
|  | High | 310 | 21 | 6.8% | 88 (29) | 65 (41) | 0.34 (0.12 - 0.90) | 66.4% | 4.50% | 23 |
| Latinx | | | | | | | | | | |
| MI | Low | 332 | 12 | 3.6% | 82 (36) | 71 (37) | 0.49 (0.16 - 1.54) | 50.3% | 1.83% | 55 |
|  | Intermediate^‡^ | 991 | 40 | 4.0% | 79 (32) | 70 (45) | 0.76 (0.44 - 1.30) | 24.3% | 0.98% | 103 |
|  | High | 323 | 13 | 4.0% | 86 (34) | 68 (40) | 0.83 (0.34 - 2.01) | 16.9% | 0.68% | 147 |
| MACE | Low | 332 | 20 | 6.0% | 82 (36) | 71 (37) | 0.48 (0.19 - 1.21) | 52.0% | 3.13% | 32 |
|  | Intermediate^‡^ | 991 | 68 | 6.9% | 79 (32) | 70 (45) | 0.79 (0.52 - 1.20) | 20.6% | 1.41% | 71 |
|  | High | 323 | 22 | 6.8% | 86 (34) | 68 (40) | 0.63 (0.30 - 1.33) | 37.4% | 2.55% | 40 |

*Major adverse cardiovascular events include myocardial infarction (fatal + nonfatal), ischemic stroke (fatal + nonfatal), and other cardiovascular death

†RRR calculated as (1-HR) X 100

‡Intermediate polygenic risk score group defined as polygenic risk quintiles 2-4

ARR, absolute risk reduction; CI, confidence interval; HR, hazard ratio; IQR, interquartile range; LDL-C, low-density lipoprotein cholesterol; MACE, major adverse cardiovascular events; MI, myocardial infarction; NA, not applicable; NNT, number needed to treat; PRS, polygenic risk score; RRR, relative risk reduction

**Supplemental Figure 1. Statin effectiveness against incident cardiovascular outcomes in self-identified White participants.** Statins users were found to have significantly higher event-free survival compared to matched statin nonusers for both incident myocardial infarction (A) and major adverse cardiovascular events (B). Major adverse cardiovascular events include myocardial infarction (fatal + nonfatal), ischemic stroke (fatal + nonfatal), and other cardiovascular death.
CI, confidence interval; HR, Hazard Ratio

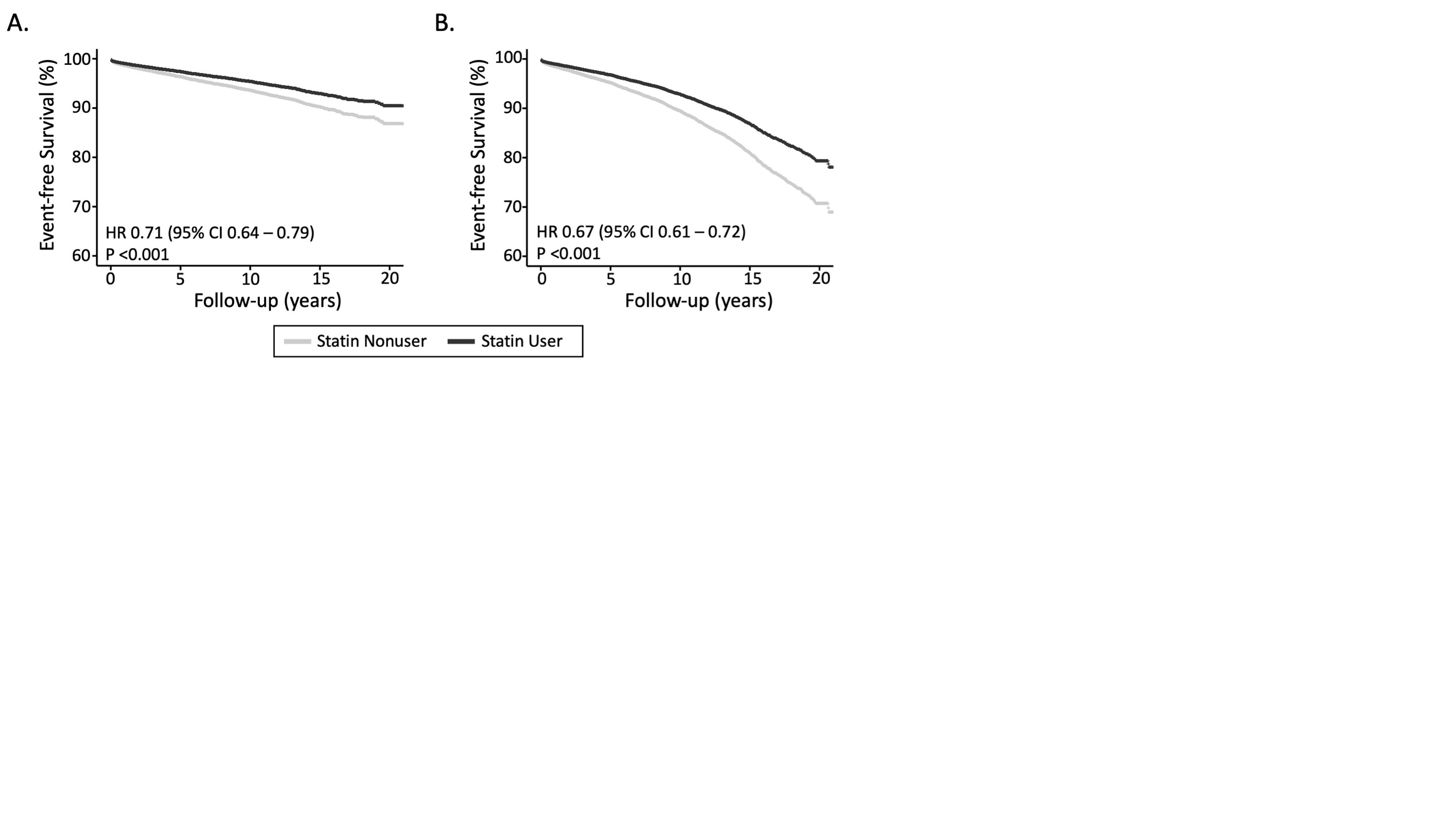

**Supplemental Figure 2. Risk of incident major adverse cardiovascular events across coronary heart disease polygenic risk score quintiles in self-identified White statin nonusers.** Higher polygenic risk score was associated with an increasing risk gradient for incident major adverse cardiovascular events. Major adverse cardiovascular events included myocardial infarction (fatal + nonfatal), ischemic stroke (fatal + nonfatal), and other cardiovascular death.
CI, confidence interval; HR, hazard ratio; PRS, polygenic risk score

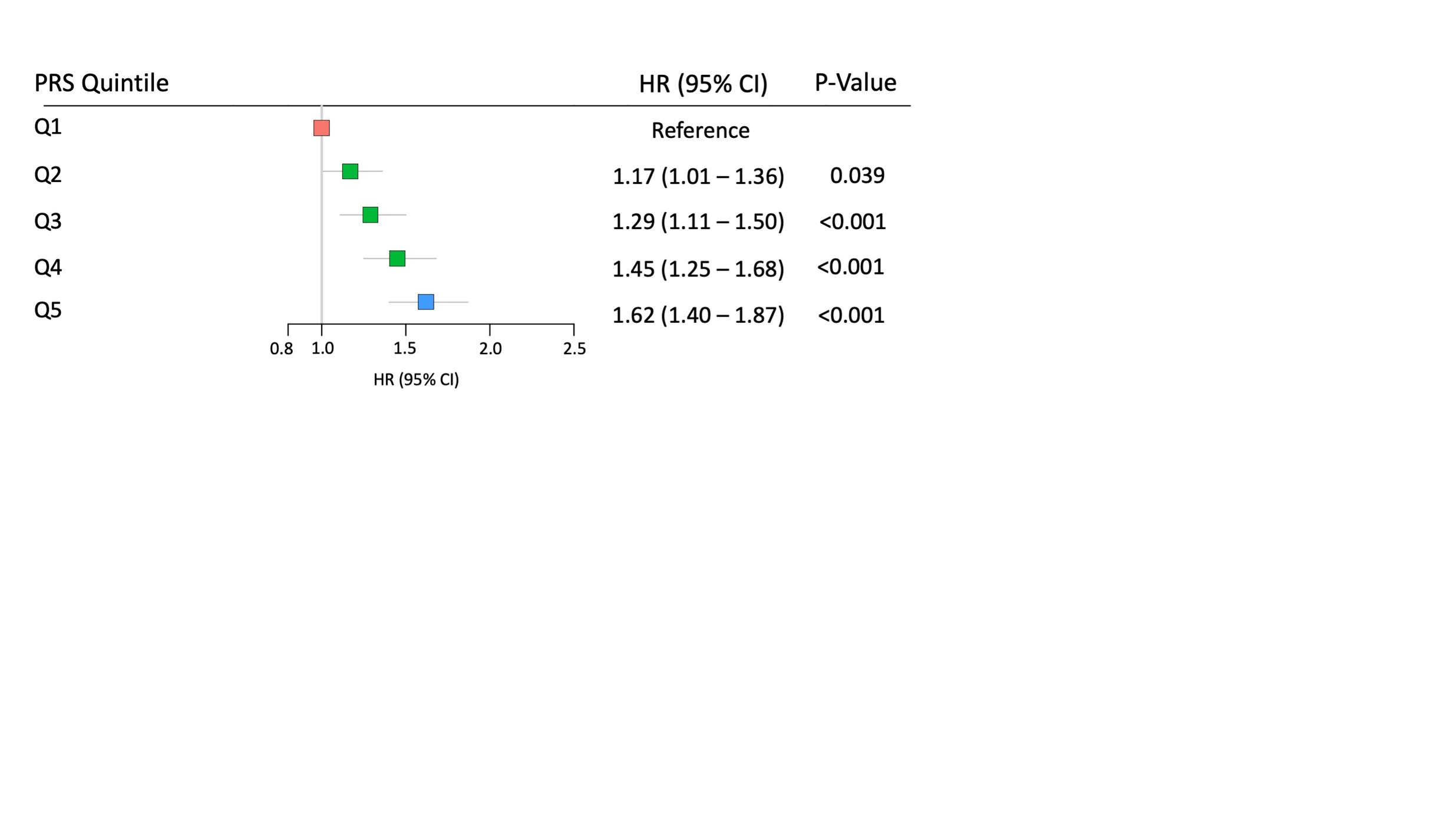

**Supplemental Figure 3. Statin effectiveness on incident major adverse cardiovascular events across coronary heart disease polygenic risk score groups in self-identified White participants.** Cox proportional hazard models of time-to-event were generated for statin users and matched nonusers to determine statin effectiveness in reducing incident major adverse cardiovascular events (defined as fatal + nonfatal myocardial infarction, fatal + nonfatal ischemic stroke, and other cardiovascular death). Increasing polygenic risk score was associated with increased statin effectiveness such that participants in the low polygenic risk score group (quintile 1) received the smallest benefit and participants in the high polygenic risk score group (quintile 5) experienced the largest benefit. Participants with polygenic risk scores in quintiles 2-4 made up the intermediate polygenic risk score group.
CI, confidence interval; HR, hazard ratio; PRS, polygenic risk score

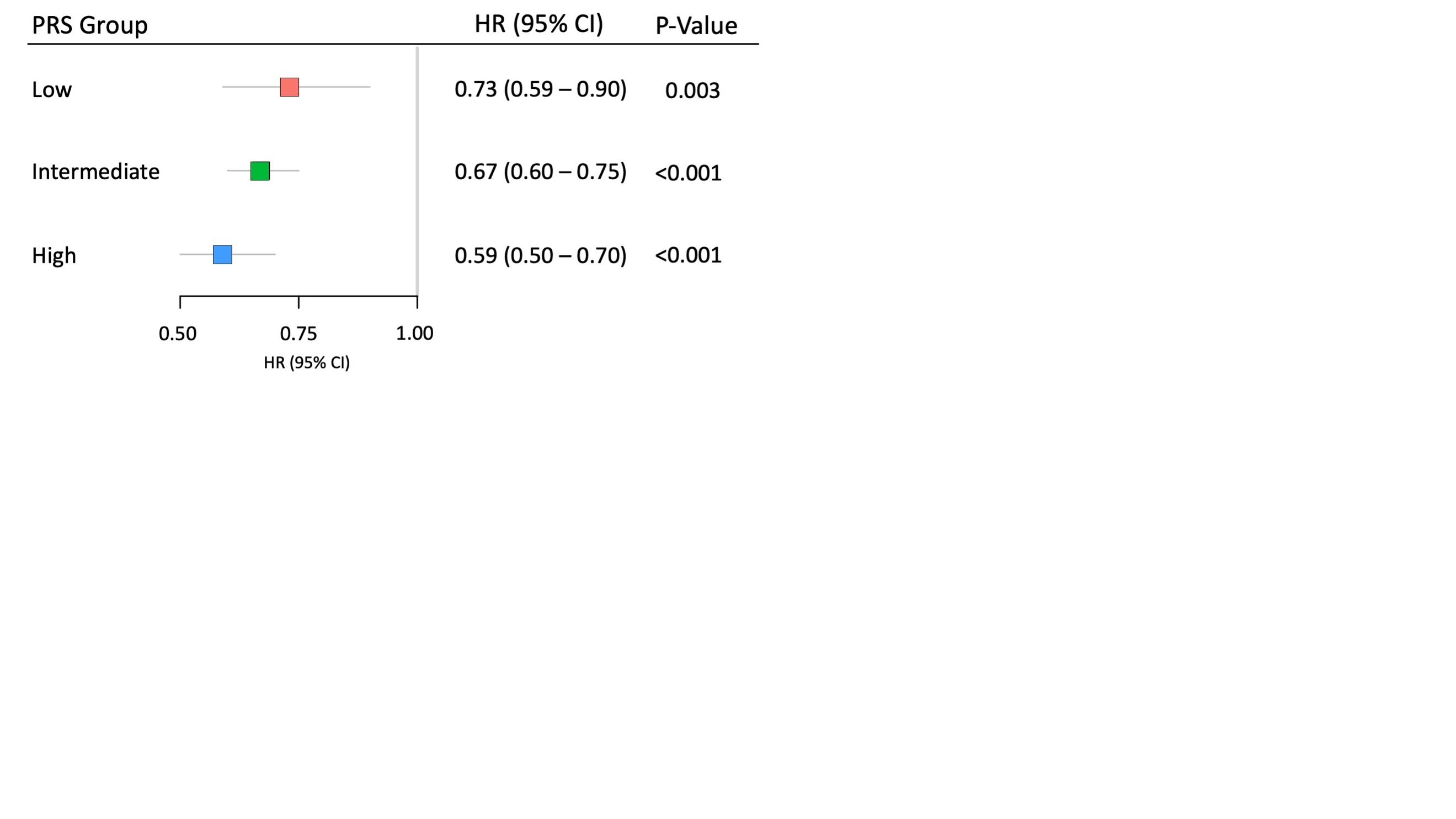

**Supplemental Figure 4. Coronary heart disease polygenic risk score distribution by self-identified race/ethnicity.**

The copy number of each variant allele (or allelic dosage for imputed variants) was weighted using the log of its corresponding odds ratio (based on published genome wide association studies linking the variant with coronary heart disease in White participants) and then summed to calculate individual polygenic risk scores for self-identified White (A), Black (B), East Asian (C), and Latinx participants (D).

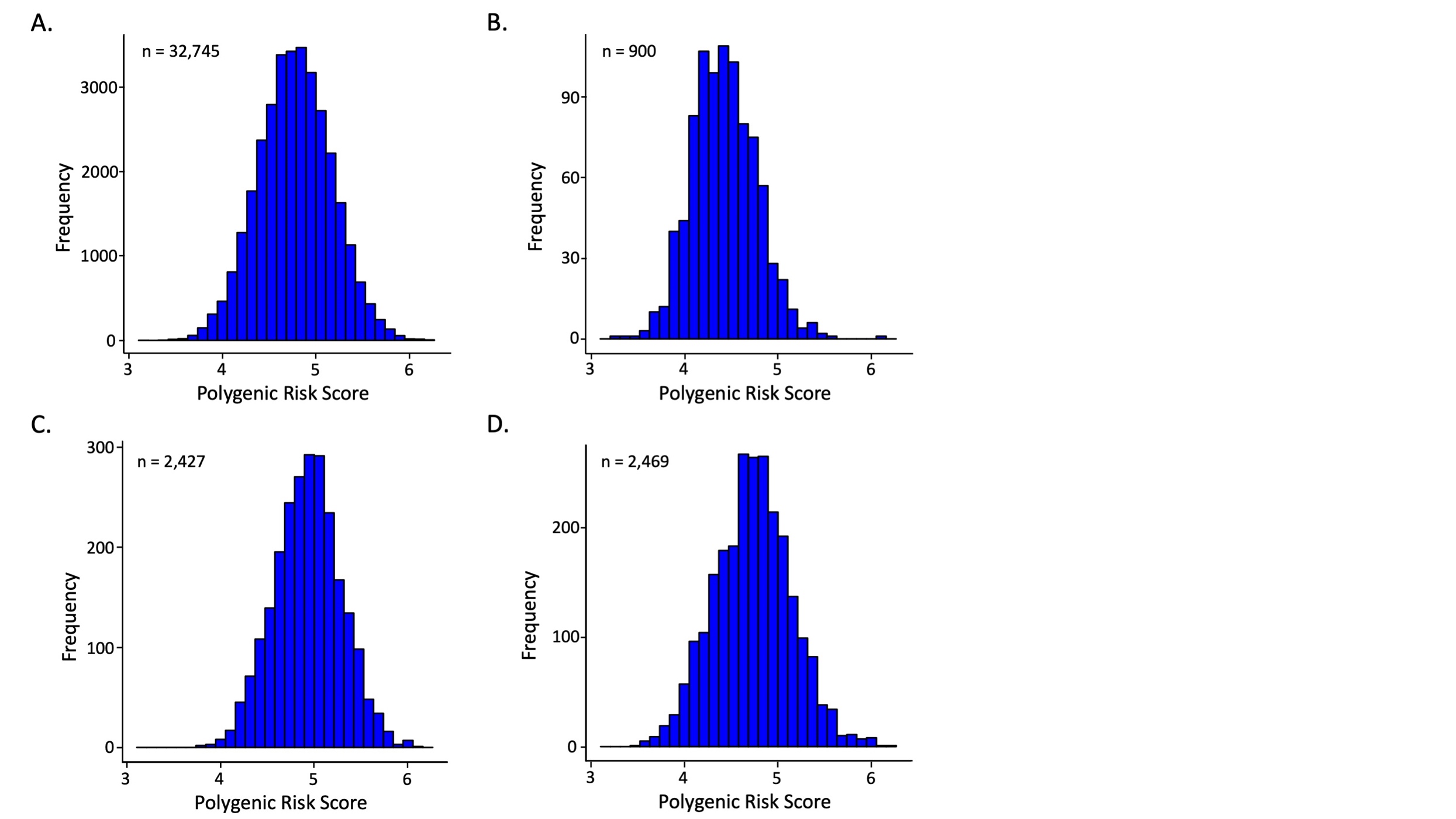

**Supplemental Figure 5. Risk of incident myocardial infarction across coronary heart disease polygenic risk score quintiles in self-identified Black statin nonusers.**

CI, confidence interval; HR, hazard ratio; PRS, polygenic risk score

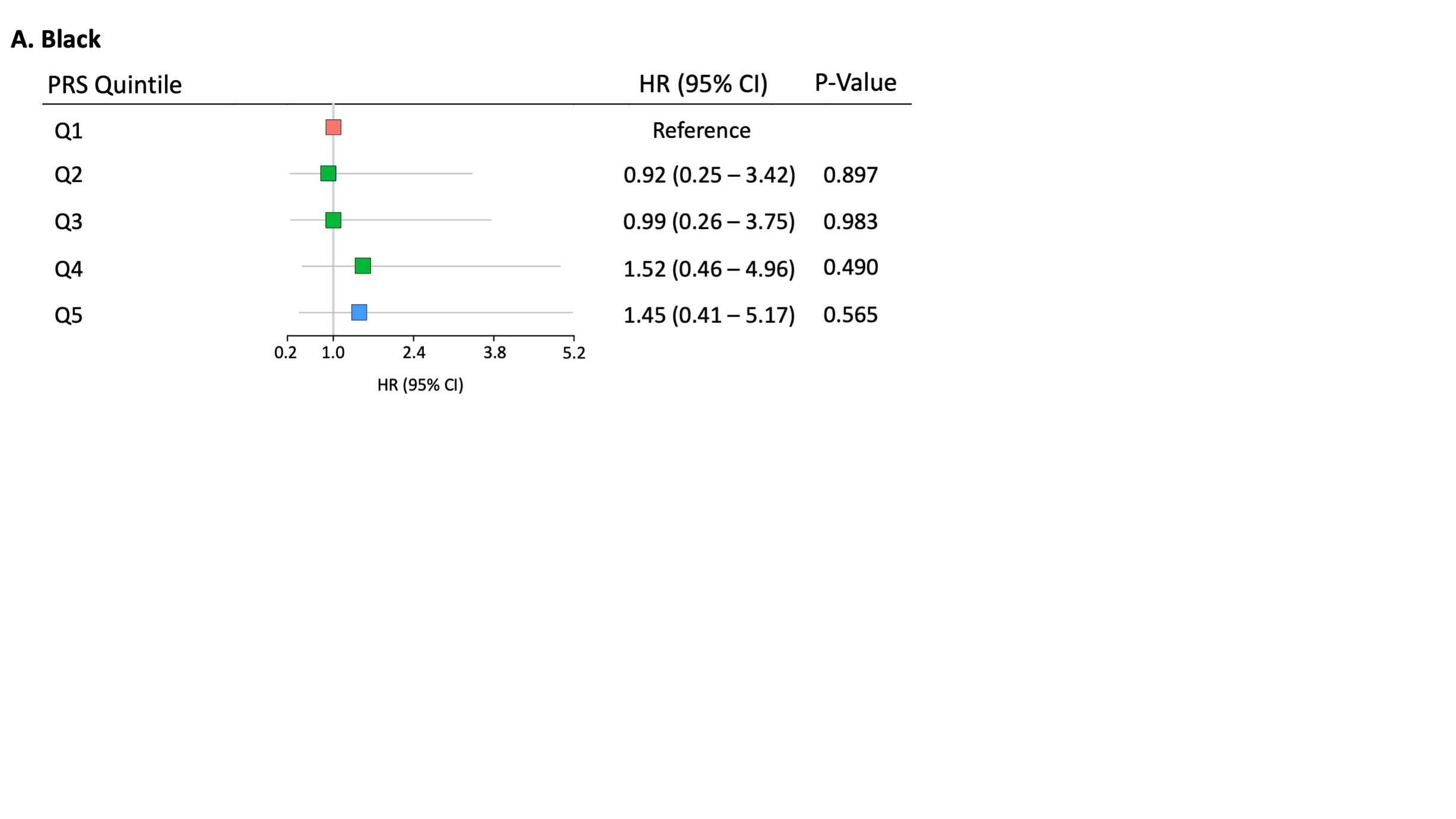

**Supplemental Figure 6. Risk of incident myocardial infarction across coronary heart disease polygenic risk score quintiles in self-identified East Asian statin nonusers.**

CI, confidence interval; HR, hazard ratio; PRS, polygenic risk score

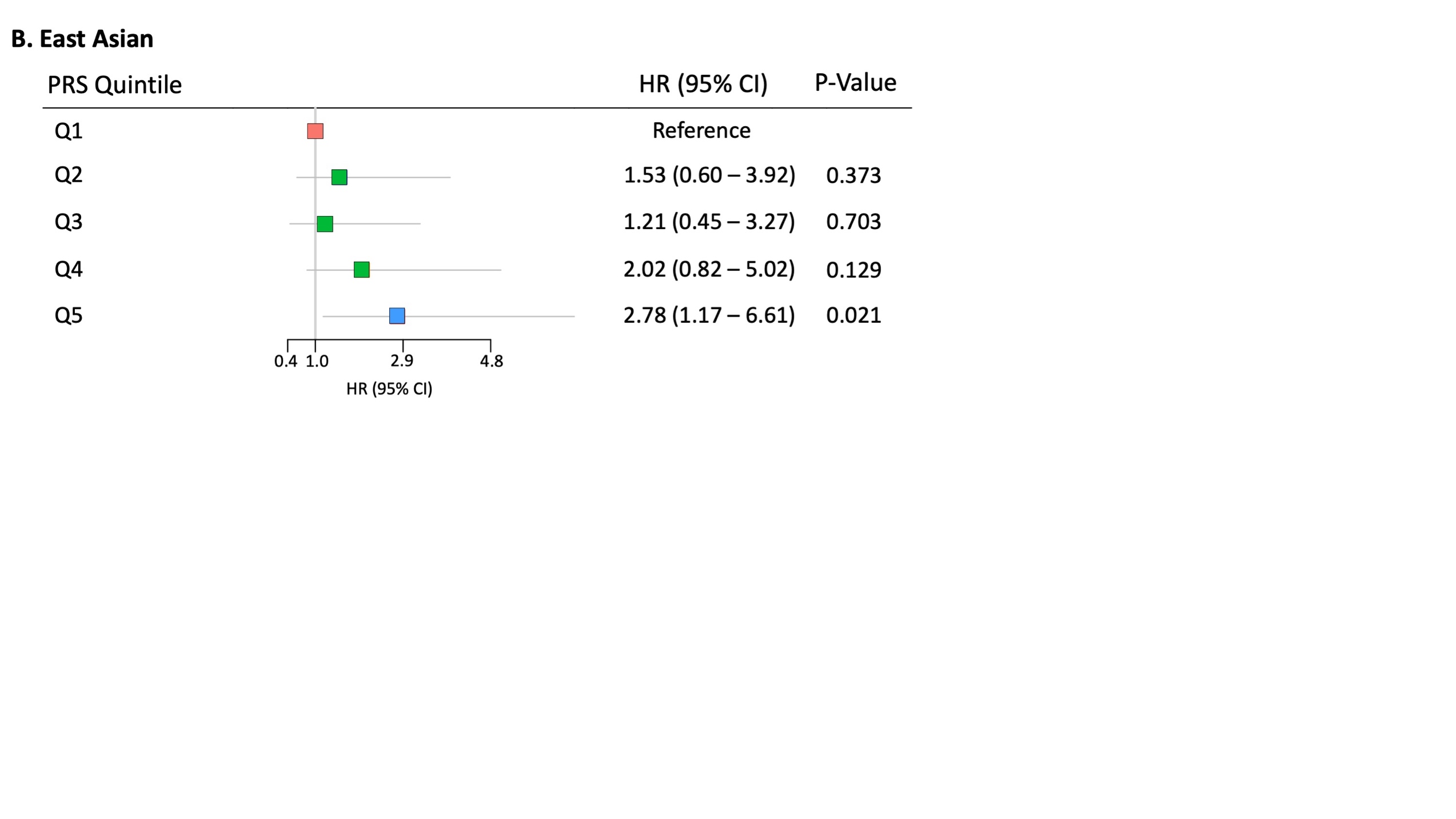

**Supplemental Figure 7. Risk of incident myocardial infarction across coronary heart disease polygenic risk score quintiles in self-identified Latinx statin nonusers.**

CI, confidence interval; HR, hazard ratio; PRS, polygenic risk score

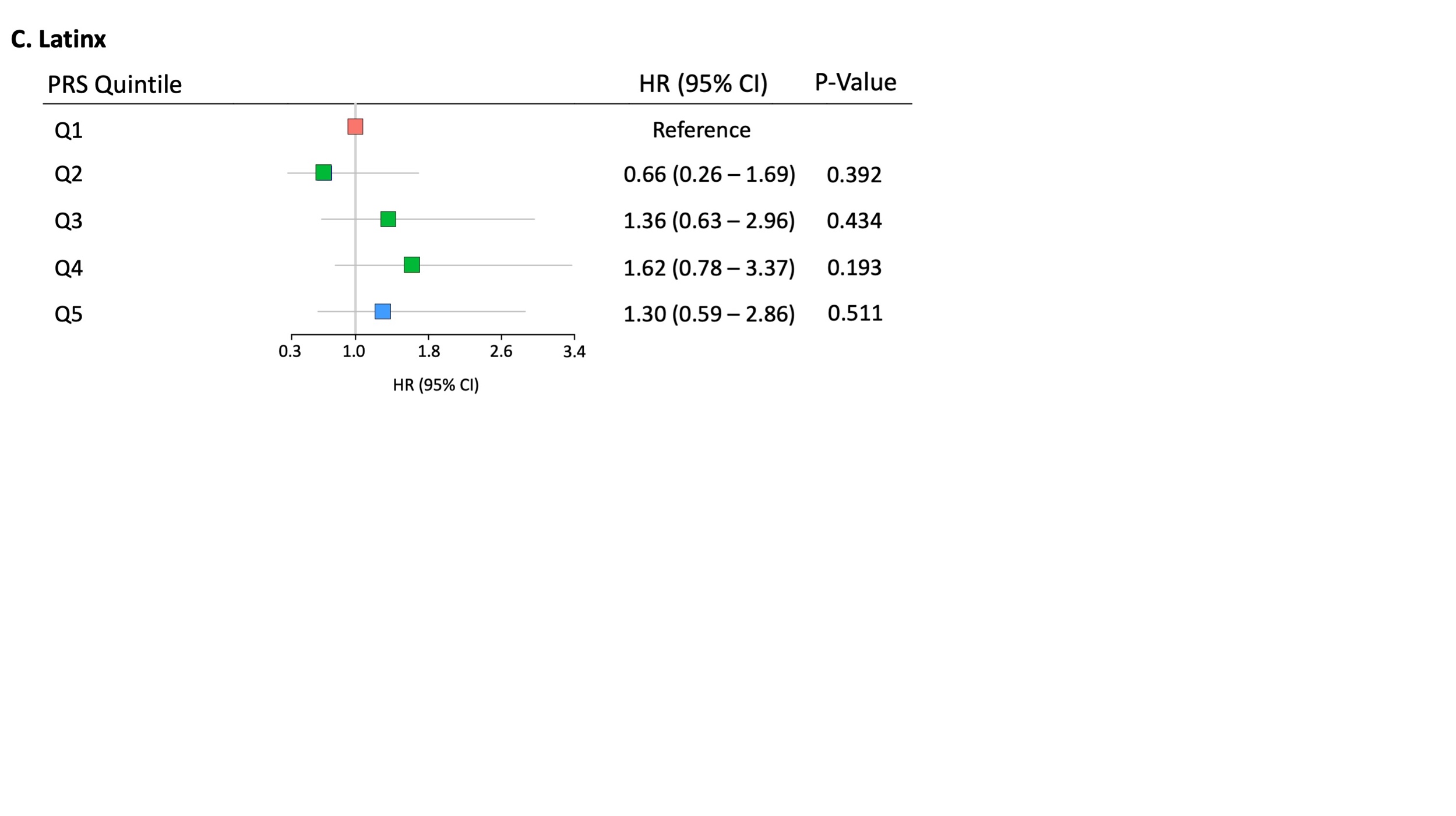

**Supplemental Figure 8. Distribution of betas for the association between coronary heart disease polygenic risk score and incident myocardial infarction in self-identified White statin nonusers at different sample sizes.**

A random subset among the full population of self-identified White statin nonusers was sampled at sample sizes of 600 (A), 1618 (B), and 1646 (C) to equal the sample size of statin nonusers in self-identified Black, East Asian, and Latinx race/ethnicity groups, respectively, before the association between polygenic risk score and time-to-incident myocardial infarction was determined. This process was repeated in bootstrap analyses (5,000 iterations) to generate betas and standard errors for each iteration. The deviation (Z-score and one-tailed p-value) of the observed beta for the association between polygenic risk score and time-to-incident myocardial infarction of each race/ethnicity group (red line) from the distribution of the resulting betas in White statin nonusers (blue bars) is displayed

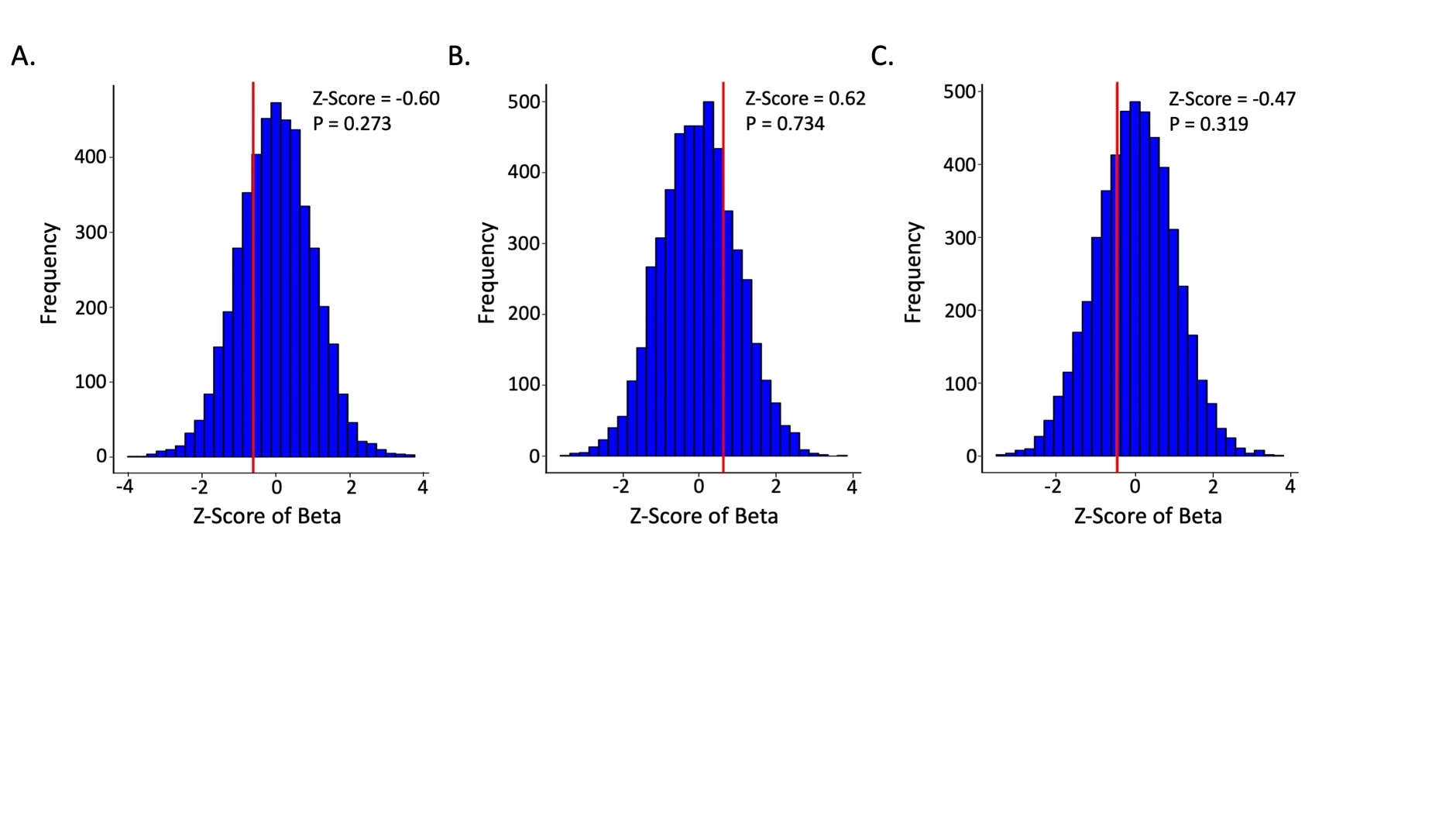

**Supplemental Figure 9. Distribution of standard errors for the association between coronary heart disease polygenic risk score and incident myocardial infarction in self-identified White statin nonusers at different sample sizes.** A random subset among the full population of self-identified White statin nonusers was sampled at sample sizes of 600 (A), 1618 (B), and 1646 (C) to match that of statin nonusers in self-identified Black, East Asian, and Latinx race/ethnicity groups. This process was repeated in bootstrap analyses (5,000 iterations) to generate betas and standard errors for each iteration. The deviation (Z-score and one-tailed p-value) of the observed standard error for the association between polygenic risk score and time-to-incident myocardial infarction of each race/ethnicity group (red line) from the distribution of standard errors in White statin users (blue bars) is displayed.
SE, standard error

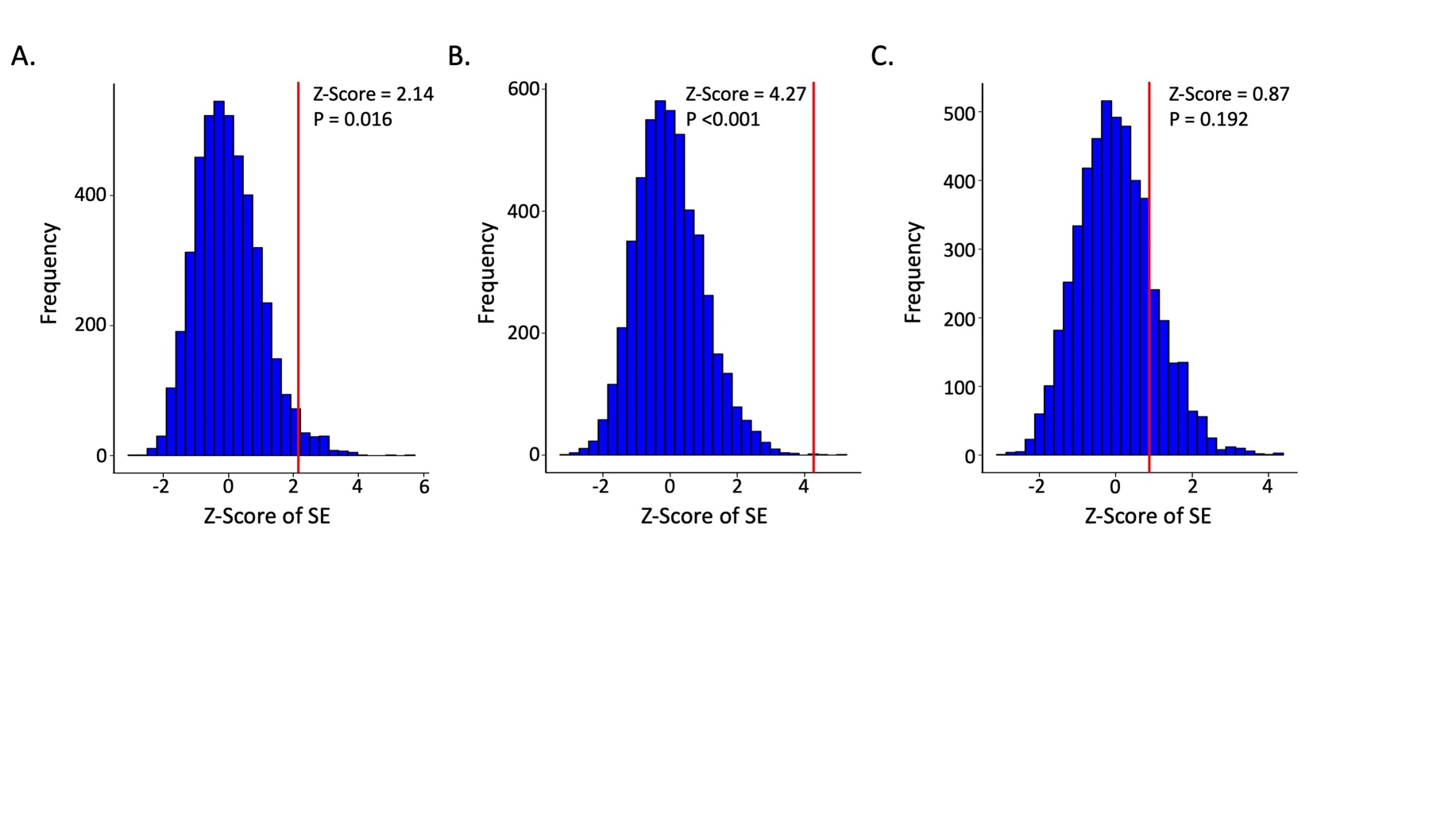
